# Post-acute COVID-19 cognitive impairment and decline uniquely associate with kynurenine pathway activation: a longitudinal observational study

**DOI:** 10.1101/2022.06.07.22276020

**Authors:** Lucette A. Cysique, David Jakabek, Sophia G. Bracken, Yasmin Allen-Davidian, Benjamin Heng, Sharron Chow, Mona Dehhaghi, Ananda Staats Pires, David R. Darley, Anthony Byrne, Chansavath Phetsouphanh, Anthony Kelleher, Gregory J. Dore, Gail V. Matthews, Gilles J Guillemin, Bruce J. Brew

## Abstract

Cognitive impairment and function post-acute mild to moderate COVID-19 are poorly understood. We report findings of 128 prospectively studied SARS-CoV-2 positive patients. Cognition and olfaction were assessed at 2-, 4- and 12-months post-diagnosis. Lung function, physical and mental health were assessed at 2-month post diagnosis. Blood cytokines, neuro-biomarkers, and kynurenine pathway (KP) metabolites were measured at 2-, 4-, 8- and 12- months. Mild to moderate cognitive impairment (demographically corrected) was present in 16%, 23%, and 26%, at 2-, 4- and 12-months post diagnosis, respectively. Overall cognitive performance mildly, but significantly (p<.001) declined. Cognitive impairment was more common in those with anosmia (p=.05), but only at 2 months. KP metabolites quinolinic acid, 3-hydroxyanthranilic acid, and kynurenine were significantly (p<.001) associated with cognitive decline. The KP as a unique biomarker offers a potential therapeutic target for COVID-19-related cognitive impairment.

---

There is evidence that SARS CoV-2 is associated with objectively tested cognitive impairment (CI) post-acute illness^1–3^ (see Table S1) but the extent, trajectory, and the nature of this new condition is unclear. Acute COVID-19 disease severity with its attendant complications is a major differential factor in the nature and extent of cognitive deficits^4^. Severe cases (cases that require hospitalization) are at risk for both indirect and direct mechanisms of COVID-19 associated brain damage, particularly from hypoxia^5, 6^, stroke, as well as the immune and inflammatory response to SARS-CoV-2^1^. Mild to moderate cases of COVID-19 (not requiring hospitalization) are still at risk of brain dysfunction, and cognitive deficits^7, 8^. Such cases provide a window into the potential mechanisms of brain injury without the confounding role of severe disease and its complications^9–11^. As most patients with SARS-CoV-2 virus do not require hospitalization, cognitive decline associated with mild to moderate COVID-19 disease would have significant implications for public health given the current global prevalence of the infection.

There are several challenges in addressing the pathogenesis of post-acute COVID-19 CI. First, there is the accurate identification and quantitation of medical co-morbidities particularly pre-morbid psychiatric conditions and illness-associated psychological distress^12^. Second, appropriate correction for normative demographic effects are imperative to accurately and specifically determine if cognitive deficits are present and to quantify their severity^13^. Many existing cognitive studies of COVID-19 that used objective testing^14^ (Table S1) have not appropriately controlled for demographics, mental health, and comorbid medical conditions. This lack of adjustment probably explains some of the large variance in the observed prevalence of CI. In addition, the previous literature on cognition in recovering COVID-19 patients has focused on patients who were hospitalized with severe acute COVID-19 disease (Table S1). Such patients can show extensive neuropsychological impairment (up to 87%) including focal insults due to stroke^3^, although improvement is detected in about half of this population at 12-months^3^. Furthermore, most cognitive studies are cross-sectional, and often have relatively small sample sizes (Table S1). Finally, studies have tended to consider olfaction and cognition separately and olfaction, in most instances, has been tested subjectively rather than objectively^15^.

To address these limitations, the current study implements a rigorous neuropsychological framework as advocated via the International Neuropsychology COVID-19 taskforce^16^ where consideration of disease severity, demographics, mental health, objectively tested olfaction, and co-morbidities is conducted a priori. Furthermore, a longitudinal study design is used because COVID-19 can have lasting effects and post-acute CI are, by definition, time-based processes. The definition of cognitive performance over time is optimized by including the initial visit overall cognitive performance, and practice effect^17, 18^. Lack of correction for these factors in cognitive studies leads to biased estimation of cognitive change^17–19^.

Using this robust neuropsychological basis, our aim was to elucidate the pathogenesis of COVID-19 associated CI by selecting a range of blood cytokines (Interferons (IFNs), major Interleukins, monocyte chemoattractant protein-1 (MCP-1), and tumor necrosis factor-α (TNF-α), peripheral biomarkers of brain injury and metabolic products. The selection of the biomarkers is founded on previous immunological findings in the same cohort, where persistently elevated IFNs (i.e., elevated expression of type I IFN (IFN-β) and type III IFN (IFN-λ1)) was associated with chest pain, and dyspnea^20^ up to 8-month post diagnosis. Next, we elected to test the components of the plasma kynurenine pathway (KP)^21, 22^ which is an IFN^23, 24^ stimulated myeloid cell mediated tryptophan degradation pathway important in immune tolerance, neurotoxicity, and vascular injury. The KP is dysregulated in COVID-19 infection^25^ including in mild cases^26^, although sex may have a moderating effect^27^. The peripheral biomarkers of brain injury have been selected because of their detectability in peripheral blood, their ability to facilitate understanding of major neurobiological mechanisms (e.g., neuro-axonal with Neurofilament Light Chain (NFL); astrogliosis with Glial fibrillary acidic protein (GFAP); blood brain barrier permeability (BBB), brain injury and astrocytosis with S100β; macrophage and granulocyte proliferation with Granulocyte macrophage colony-stimulating factor (GMCSF)), and because each is abnormally elevated in COVID-19 infection (NFL^28^; GFAP^29^; S100β^30^; GMCSF^31^).

The aims of the current study were to determine the prevalence of CI and the trajectories of overall cognitive performance (both corrected for age, education, and sex) in COVID-19 cases recovering from mostly mild-moderate acute infection across a 12-month period. This is achieved by using the widely recognized Cogstate computerized battery that is ideal for serial repeating testing. Furthermore, we aimed to elucidate the pathogenesis of the observed cognitive changes. Moreover, the clinical significance of CI was assessed against functional status. The pathogenic axes assessed included systemic disease severity (and indirectly hypoxia), lung function, objectively tested olfaction, pre-existing and illness-associated anxiety and depression, medical comorbidities, initial visit CI, blood cytokines, the KP and peripheral biomarkers of brain injury. Time-variant predictors (all biomarkers but IFN-β IFN-λ1, olfaction) were tested for their main and time association with overall cognitive performance. The KP was also tested for its trajectory and its dynamic links to other biomarkers.

Despite the exploratory angle of our study, we expected to detect a level of mild cognitive impairment, but without evidence of severe cognitive deficits or rapid deterioration based on previous studies in similar COVID-19 cases. We hypothesized that systemic disease severity, lung function, objectively tested olfaction, pre-existing and illness-associated anxiety and depression, initial visit CI, IFN-β, Quinolinic Acid as well as involvement of peripheral biomarkers of brain injury would show some level of association with cognitive decline.

## Methods

The study reporting follows the STROBE guidelines for cohort and observational studies.

### Participants

A total of 128 unvaccinated participants with nasopharyngeal swab-confirmed SARS-CoV-2 infection were enrolled through the Sydney St. Vincent’s Hospital COVID-19 ADAPT prospective study. The study is ongoing, the first exam was on May 14^th^, 2020; and the data for the current analyses were censored on September 15^th^, 2021.

### Ethics

The study protocol was approved by St. Vincent’s Hospital Human Research Ethics Committee (Reference #2020/ETH00964) and all participants signed an informed consent form.

### Design

The study has two main longitudinal components: 1. the neurocognitive assessment, and 2. the clinical and blood test visit assessment.

For the clinical and blood test visit, 128 participants completed the first assessment 2.7 (SD=1.3) months post diagnosis; 126 completed a second visit 4.8 (SD=1.8) months post diagnosis; 117 completed a third visit 9.0 (SD=2.2) months post diagnosis; and 101 completed a fourth visit 12.7 (SD=1.4) months post diagnosis. For the neurocognitive assessment, 127 completed their 1^st^ assessment 2.7 (1.2) months post diagnosis; 121 completed their second visit 5.3 (SD=2.7) months post diagnosis; and 101 completed a third visit 12.9 (SD=1.6) months post diagnosis. Participants who had concomitant neurocognitive and clinical blood tests visits were the same as those who completed the neurocognitive assessments. For clarity, these assessment times will be referred to as 2, 4 8 (only for the clinical and blood test visit) and 12 months post diagnosis when describing the time effect. However, because the time effect is unbalanced across subjects (that is every individual came at a specific time); the analyses use a continuous time effect (see data analyses section for more details).

The study also had three cross-sectional components: 1. the mental health assessment which occurred 2-months post diagnosis at the same time as the neurocognitive assessment; 2. the recording of medical (including neuropsychiatric) history which occurred when the patient was first enrolled, typically a few days/weeks before their first neurocognitive assessment. 3. the detailed recording of acute COVID- 19 illness history and the measurement of lung function. Based on acute COVID-19 illness history, the participants were divided into three acute severity groups (mild, moderate, and severe). Detailed information for this classification has been previously reported in Darley et al.,^32^. Briefly, mild acute disease patients were community-managed with minor, mostly upper respiratory symptoms. Moderate acute disease patients were community-managed with fever or chills and experienced at least one of the following symptoms: cough, shortness of breath, chest pain, nausea or vomiting, diarrhea, mild instances of altered consciousness or confusion, abnormal lung function. Patients were also classified as moderate illness if they did not experience fever or chills and presented with at least two of the aforementioned symptoms. Patients were classified as severe if they had been hospitalized. The cohort at 2-months post diagnosis was composed of 39% with mild disease (n = 50), 52% with moderate disease (n = 66), and 9% with severe disease (n = 12) (see ^32^ for details).

### Procedures

#### Neurocognitive testing

The neurocognitive test selection was informed by the International Neuropsychology COVID-19 taskforce^16^. The CogState Computerized Battery (CCB) is a widely used and validated cognitive screening test that is culturally fair. The CCB version used in this study included Detection (reaction time), Identification (reaction time), One Card Learning (accuracy), and One Back (reaction time and accuracy). The tasks’ selection was based on the recommendations by the NeuroCOVID-19 International Neuropsychology Taskforce^16^. Specifically, the tasks target cognitive abilities affected by acute respiratory illnesses, immune compromise, and cerebrovascular diseases, thus covering the main mechanism of disease development of COVID-19^16^. Additionally, the CCB has normative data, including a sub-set specifically validated for the Australian population that has similar demographics as the current study population^18^. Further, the CCB is ideal for repeated testing (test-retest reliability r = 0.8)^18^. For the current study LAC trained provisional neuro/psychologists and nurses in the administration of the CCB following our previously published protocol^33^.

The CCB reaction time data was log-transformed; while the accuracy data was arcsine root-transformed to approximate the normal distribution. We then applied age, sex and education corrections to the log or arcsine root transformed data using the Australian normative data. This produced five demographically corrected z-scores. Then, the five z-scores were averaged into a mean z-score for each individual where a higher z-score represents a better global performance. The rationale for summarizing the CCB performance into a global composite score is twofold: 1. this composite score is more reliable than any of the individual measures^18^; and 2. the composite score is the most sensitive and specific to clinically significant cognitive impairment^34^. Next, the follow-up (4 and 12 months) data were corrected for task-specific practice effects using published norms from the demographically comparable Australian healthy controls^18^.

To classify clinically relevant global CI, the five individual z-scores were transformed into deficit scores ranging from 0 to 5. A deficit score of 0 indicated no impairment (z ≥ −1.0), 1 indicated mild impairment (z < −1.0 to −1.5), 2 indicated mild to moderate impairment (z < −1.5 to −2.0), 3 indicated moderate impairment (z < −2.0 to −2.5), 4 indicated moderate to severe impairment (z < −2.5 to −3.0), and 5 indicated severe impairment (z < −3.0)^34^. The deficit scores were subsequently averaged into a Global Deficit Score (GDS) where a higher GDS indicated greater impairment. We used best-evidence clinical cut-off (GDS>.05) to classify at least mild overall CI^34^.

The NIH Toolbox Odor Identification Test (NIH OIT) is a validated screening test which assessed olfaction performance. The participants were requested to smell nine scented scratch-and-sniff cards sequentially and match the card’s smell to one of four depictions of different odors on an iPad. The test included a variety of odors of food and objects with which people commonly interact in their environment^35^. The NIH OIT has been validated against more comprehensive, and well-established olfactory tests (the University of Pennsylvania Smell Identification Test and the Brief Smell Identification Test) and comes with a large set of normative data corrected for age, education, sex, and race/ethnicity^35^. The NIH OIT raw score reflects the number of odors correctly identified (0-9). This raw score was transformed into demographically corrected T-scores within the NIH Toolbox based on the norms that are associated with the application. For the NIH OIT data, the demographically corrected T-scores was classified as “impaired” if the T-score was <40, as per convention to identify at least mild olfactory impairment^35^.

#### COVID-19 Associated Functional Status

At 2- and 4-month post diagnosis, the participants completed a COVID-19 functional status scale answering the four following statements: I have fully recovered after COVID-19; I feel confident returning to my pre-COVID work; I have returned to my usual activities of daily living; I have returned to my normal exercise level. For each statement, the participant rated whether they strongly agree, agree, slightly agree, slightly disagree, disagree, or strongly disagree, rated from 1 to 6.

#### Mental Health Assessment

Mental health was assessed with three screening tools to capture anxio-depressive symptoms at the first assessment, 2 months post diagnosis. In addition, participants reported whether they had been formally diagnosed with any psychiatric conditions. We used the Depression in the Medically Ill (DMI-10) which is a 10-item questionnaire. The DMI-10 accounts for confounding factors of the physical illness by focusing on cognitive symptoms of depression and avoiding measuring physical symptoms common to both depression and physical illnesses (e.g., changes in sleep, appetite, and body weight). DMI-10 scores range from 0 to 30 and a score of 9 or greater reflects a possible major depressive episode. The Impact of Event Scale Revised (IES-R) is a 22-item questionnaire, which measured post-traumatic stress disorder (PTSD) symptoms of the past 7 days. The questionnaire consisted of three sub-scales: intrusions, avoidance, and hyperarousal. Patients indicated their level of agreement to statements about commonly experienced PTSD symptoms on a 5-point scale ranging from “not at all” to “extremely”. IES-R scores range from 0 to 88 and a score of 33 or above indicates a likely PTSD diagnosis. The Anxiety-Depression Subscale of the Somatic and Psychologic Health Report (SPHERE-34) was used to assess anxio-depressive symptoms. Participants used a 3-point scale (from “never or some of the time” to “a good part of the time”) to indicate the frequency of experiencing common depression and anxiety symptoms over the past few weeks. Scores on the anxiety-depression subscale ranges from 0 to 14 and a score of 2 or more indicates a clinically significant mood disorder. The total scores of the three mental health questionnaires were summarized by Principal Component analysis as total scales’ scores were highly correlated (r>.80). We kept the first component which explained 80% of the mental health data’s variance and a higher score indicated greater mental health symptoms as having a pre-existing mental health condition was highly associated with the mental health composite score (p<.001).

#### Medical visit, blood draw and assessment of lung function

Patients underwent a demographic and medical history inventory 2-months post- diagnosis. Patients reported their age, sex, geographical background, and self-identified race/ethnicity (country born; whether of English-speaking background, white, black, Hispanic, other), level of education and medical co- morbidities, including pre-existing neurological and psychiatric conditions. In addition, patients’ acute symptom severity was classified based on medical data (e.g., lung function assessments, evidence of hospitalization and ICU stay) and patient’s self-report of acute symptoms. The detailed lung function procedures have been published in Darley et al., 2021^32^. For the current analyses, we selected the two outcomes that were most related to disease severity in the 2021 study analyses: at 2-month post diagnosis % predicted Total Lung Capacity (TLC) and % predicted Haemoglobin-Corrected Diffusing Capacity of the Lung for Carbon Monoxide (DLCO). In the current study severe cases had lower % predicted TLC (p<.05) compared to mild and moderate cases, which did not differ from one another. Further, severe cases had lower % DLCO (p<.05) compared to both mild and moderate cases. Moderate cases had lower % predicted DLCO compared to mild cases although this was not significant.

#### Quantification of the KP metabolites using Ultra High Performance Liquid Chromatography (uHPLC)

Approximately 180 µL of patient serum was deproteinized by adding equal volume of with 10% (w/v) trichloroacetic acid. After, the mixture was centrifugated at 4°C for 10 min at 12,000 rpm and filtered using 0.20μm polytetrafluoroethylene syringe filters (Millex, Merck) into a new analyzer vial. The KP metabolites were analyzed using an Agilent 1290 series uHPLC system that includes a temperature-controlled column compartment (40° C) and auto- sampler (4° C), diode array (G4212A; Agilent) and fluorescence detector (G1321B; xenon flash lamp; Agilent). The mobile phase was 0.1 M sodium acetate at pH 4.6 and the assay was run with an isocratic flow rate of 0.75ml/min in an ZORBAX Rapid Resolution High Definition C18 column (2.1 mm x 150 mm, 1.8 μm particle size; Agilent) for 12 min. The fluorescence detector was set at excitation/emission wavelength of 280nm/438nm for detection of tryptophan (TRP) and 320 nm/438 nm for detection of 3-hydroxyanthranilic acid (3HAA) and anthranilic acid (AA). Kynurenine (KYN) and 3-hydroxykynurenine (3HK) were detected using a UV detector set to measure absorbance at 365 nm (reference signal off). The results were calculated by interpolation using a six-point calibration curve and expressed as μmol/L or nmol/L. The chromatogram output of KP metabolites was analyzed using the Agilent OpenLAB CDS ChemStation (Edition C.01.04).

#### Quantification of the KP metabolites by Gas Chromatography/Mass Spectrometry (GC/MS)

Agilent 7890 A GC coupled with Agilent 5975 C MS detector and a DB-5MS column (0.25 mM film thickness, 0.25 mm x 30 m capillary column) was used to quantify picolinic acid (PIC) and quinolinic acid (QUIN). Fifty µL of deproteinized serum used in uHPLC was mixed with deuterated internal standards respective to the metabolites of interest were derivatized for quantification. For the sample derivatization the deproteinized samples and deuterated internal standards were dried under vacuum and derivatized with trifluoroacetic anhydride for 10 min at 60°C followed by 1,1,1,3,3,3hexafluoroisopropanol for extra 10 min at 60°C. Fluorinated esters were then extracted into toluene and washed with 5% sodium bicarbonate. The upper organic layer was collected and washed with 1 mL MilliQ water, and dried using sodium sulphate packed pipette tips. Samples were then transferred into a new analyzer vial for analysis and injected under a splitless mode onto the GC capillary column. The analysis was carried out with the MS operating in negative chemical ionization mode. Selected ions (m/z 273 for PIC, m/z 277 for 4-PIC, m/z 467 for QUIN and m/z 470 for d3- QUIN) were simultaneously monitored. GC oven settings were as follows: oven temperature was held at 75°C for 3 min and then ramped to 290°C at a rate of 25°C/min and held at 290°C for 4 min for a total run time of 15.6 min. A series of mixed non-deuterated and deuterated standards of PIC and QUIN were used for a six-point standard curve to interpolate the quantity of the sample readout. Levels of PIC and QUIN were calculated and expressed as nmol/L. The chromatogram output of PIC and QUIN was analyzed using Agilent GC/MSD ChemStation software (Edition 02.02.1431).

#### Quantification of IFN-β and IFN-λ1

IFN-β and IFN-λ1 were quantified via LEGENDplex Human Anti-Virus Response Panel as previously described^20^.

##### Cytokines and chemokines assay

The quantification of the cytokines and chemokines in COVID-19 patients’ sera was carried out by Eve Technologies (Calgary, Canada) using the human focused 15-Plex Discovery Assay. The cytokines and chemokines that were quantified include GMCSF, IFN-γ, Interleukin-1beta (IL-1β), IL-1 receptor antagonist (IL-1 Ra), IL-2, IL-4, IL-5, IL-6, IL-8, IL- 10, IL-12p40, IL-12p70, IL-13, MCP-1, and TNF-α.

##### Enzyme-linked immunosorbent assay

Human GFAP and S100B enzyme-linked immunosorbent assay kits were purchased from R&D Systems (Minneapolis, MN, USA), and NF-L enzyme-linked immunosorbent assay kit was purchased from Novus Biologicals (Centennial, CO, USA). The measurement of these proteins in the sera of the COVID-19 patients was carried out according to manufacturer’s instructions and the concentrations of GFAP, NF-L, and S100b were calculated based on the optical density of the samples to the standard curve.

### Statistical analyses

#### Missing data

There were no missing data for cognition or the KP data. IFN-β and IFN-λ1 were measured concomitantly to the 2-month post diagnosis cognitive session in a representative sub-sample of n=68^20^. Other cytokines, inflammatory biomarkers and peripheral biomarkers of brain injury had no missing data and the lowest plausible value was imputed when the product was below detection (See Table S2 in supplemental file). One case had pre-morbid anosmia and thus no smell data was collected. Eight cases did not have follow-up olfaction data at 12 months due to technical issues. The attrition in this with a neurocognitive assessment (loss to follow-up and/or not yet completed follow-up) was relatively small, thus we did not conduct any sophisticated missing data analyses; and cases who remained in the study did not significantly differ in any characteristics compared to those who were not assessed at follow- up.

To present the demographic and clinical characteristics of the study sample in detail, we elected to present the data for the entire sample and by COVID-19 severity subgroups at the visit closest to acute infection (2-month post diagnosis). Comparisons between the subgroups were conducted with ANOVA, Chi-Square as appropriate.

Next, we used a series of multilevel models (i.e., general linear mixed models) to test which factors contributed to cognitive change over the entire study period. We sought to target an optimal balance between parsimony and good fit. Residual maximum likelihood (REML) was used in estimating and reporting all model parameters. Denominator degrees of freedom were estimated using the Satterthwaite method.

#### Time effect and model’s analytic framework

Graphically, time effect is represented by visit time, that is 2, 4, 8 (for the KP), and 12 months post diagnosis. Depending on the models’ aim, time is either used as visit time or centered so that the first visit becomes the reference. When relevant, time is also used as a continuous fixed effect and/or random nested effect as the exact month (2 decimals) post diagnosis for each participant. Continuous time was also centered using the sample’s mean. Polynomial time effects (square, quadratic and cubic) were tested and kept in the models when they significantly improve the model’s fit. The significance of new fixed effects was evaluated with univariate and multivariate Wald tests. The significance of the random effect was evaluated using -2LL ratio.

We followed the analytic steps recommended by Hoffman L^36^. We first tested the within and between variance pattern of the longitudinal data (neurocognition or KP outcome) using a means saturated model by REML estimation and an unstructured covariance type, and time as centered visits. We then determined whether the addition of a random subject effect and then a random time slope significantly improved the model. At this stage, we also included a model’s personality for the intra-individual variance. Several models’ personality was tested using the -2LL ratio (smaller is better). Identity and diagonal repeated covariance matrices fitted our data best. At this stage, time was used as a centered continuous time effect. Next, when relevant, we tested whether including polynomials time effects improved the model’s fit. Then, we also tested whether the addition of a random quadratic centered continuous time effect improved the model. Finally, we tested a sex effect (male=1; female=2) and its interaction with time. For each model, the time fixed effect was tested using a Wald test (set at p<.05 for significance). Random effects were tested using a -2LL ratio (smaller is better). Overall model fit was tested using the AICc (smaller is better).

#### Cognitive analyses

The cognitive data was normally distributed. To assess any association between olfaction and cognition, we first tested whether they were associated at 2-months post diagnosis using Person correlations and t-test and Chi-square as relevant. Next, using standard clinical cut- offs for cognition, we extracted prevalence of impaired performance across the study period and compared them with Chi-Square. Furthermore, to assess the longitudinal profile of the cognitive data, we used the demographically and practice corrected CCB mean z-score (z-scores have a mean of 0 and a SD of 1) as our primary outcome. Sex, age, and education were not added as these were already corrected for. Next, based on prior knowledge^37^, we added 2-month post diagnosis CI status (initial visit impaired versus unimpaired status) as a fixed effect predictor and its interaction with time. Using the best fit model, we tested olfaction, mental health, medical comorbidities, lung function, and disease severity as fixed effect and their time interaction were significantly associated with the outcome.

#### Functional status and cognitive impairment

To assess any association between functional status and cognitive impairment, we compared the participants classified as having mild to moderate cognitive impairment (as defined above) versus those who were classified as cognitively normal using t-tests. We used the individual functional status responses, as those corresponded to different aspect of everyday function. Cognitive impairment was corrected for demographics and corrected for practice effect at 4-month post diagnosis.

#### KP analyses

Prior to analyses, the KP data were inspected for outliers and data were winsorized in 7 instances to minimize the effects of these outliers. To approximate the Normal Distribution, the data was Log transformed for PIC, QUIN, 3HK, AA, and 3HAA. A square root (√) transformation was applied to KYN. Because the KYN showed somewhat binomial distribution which only slightly improved with the square root tranformation, we also dummy coded the KYN as 0=within normal range, and 1=KYN >3uM (cut-off based on aged norms^32, 33^). The raw TRP was normally distributed. To illustrate the KP intra-dynamic, correlation between the KP metabolites was assessed for the 2-month data. The KP longitudinal dynamics were tested in each metabolite separately. Using the best fit models, we tested whether mental health, medical comorbidities, lung function, disease severity and sex as fixed effect. Their time interactions were significantly associated with the outcome.

#### Cognition and KP profile

We used the final cognitive models to start with and removed polynomials and random effects if the model did not converge. Each KP metabolite was entered as a fixed continuous predictor and its interaction with continuous time. Fixed effects were added if they had significantly improved the model’s fit in the previous steps.

#### The KP, IFN-β and IFN-λ1 analyses

IFN-β and IFN-λ1 were log transformed to approximate the Normal Distribution. Using a sub-sample (n=62) of the current cohort which had IFN-β and IFN-λ1 measured concomitantly to the 2-month post diagnosis cognitive session, we determined their association with the longitudinal KP metabolites^20^. To do this, we conducted a mixed effect model where the longitudinal KP metabolites data were entered as outcomes (separately tested), subject as a random effect, persistent lung symptoms (n=31) versus no lung symptoms (N=31) as a fixed main effect, the IFN as a fixed main effect and its interaction. We included the persistent/no lung symptoms as a fixed effect because this sub-sample had been purposely selected with this in mind^20^. Persistent lung symptoms were defined as persistent chest pain and/or dyspnoea at 2- and 4 months post diagnosis^20^.

#### KP, cognition and additional cytokines (IFN-γ, Interleukins, MCP-1, TNF-α) and peripheral brain biomarkers (NFL, S100β, GFAP and GMCSF) analyses

All biomarkers were Log transformed except for MCP-1 which was normally distributed. We conducted final mixed effect models using two of the KP metabolites (top and low sections of the KP: √KYN and Log QUIN) that were significantly associated with cognition in the previous analyses as outcomes. We also tested the biomarkers directly against cognition. For each model, we tested main effects and time interaction effects as in previous models.

Analyses were conducted in SPSS V.28 and JMP V.15 (SAS Inc). Considering the exploratory nature of the analyses and the need to control for experiment-wise comparisons’ error rate, we used False Discovery Rate (FDR) correction^38^ within each main model. Moreover, effect sizes were extracted to assist in the interpretation of the significance of the effects.

## Results

### Sample Characteristics

The group is composed of 42% women. Patients are aged between 20 and 79 years old with an average age of 46.6 years old. The acute severe subgroup is older and composed of more males than the two other subgroups (Table 1).

**Table 1:** Demographic, comorbidity, and mental health characteristics <u>at 2**-m**onths post diagnosis</u> by acute disease severity

| Sample Characteristics | Full Sample<br>(n=128) |  | Mild (n=50) |  | Moderate (n=66) |  | Severe (n=12) |  | *p* | *Comparisons<sup>2</sup>* |
| --- | --- | --- | --- | --- | --- | --- | --- | --- | --- | --- |
|  | *M* | *SD* | *M* | *SD* | *M* | *SD* | *M* | *SD* |  |  |
| **Age (Years)** | 46.7 | 15.0 | 48.7 | 15.5 | 42.8 | 14.0 | 59.3 | 9.1 | .0007 | Severe > Mild & Moderate<br>(p<.03) |
| **Educations (Years)** | 15.3 | 2.8 | 15.6 | 2.8 | 15.1 | 2.7 | 14.9 | 3.6 | .54 | ns |
| **Mental Health Composite** | 0.1 | 1.6 | -0.1 | 1.7 | .2 | 1.6 | -0.1 | 1.3 | .46 | ns |
| **DMI-10** | 6.1 | 6.6 | 5.6 | 6.1 | 6.7 | 7.1 | 5.1 | 5.8 | .45 | ns |
| **IES-R** | 12.8 | 14.1 | 10.8 | 15.0 | 13.4 | 12.7 | 18.1 | 17.0 | .15 | ns |
| **SPHERE** | 1.9 | 2.6 | 1.8 | 2.9 | 2.2 | 2.6 | 0.5 | 0.9 | .16 | ns |
|  | *n* | % | *n* | % | *n* | % | *n* | % |  |  |
| **Sex (Female)** | 53 | 41.7 | 20 | 15.7 | 32 | 25.2 | 1 | 0.8 | .03 | Severe < Mild & Moderate<br>(p<.03) |
| **ESB<sup>1</sup>** | 115 | 90.5 | 45 | 91.2 | 60 | 90.9 | 10 | 83.3 | .66 | ns |
| **Medical Comorbidities<sup>3</sup>** | 46 | 36.2 | 16 | 12.6 | 22 | 17.3 | 8 | 6.3 | .07 | Mild & Moderate > Severe<br>(p=.03) |

<sup>1</sup> ESB: English speaking background assessing geographical background/race/ethnicity. People of non-ESB included Asian Australians born in Australia for the majority, and then South Americans, non-English speaking European people; all were fluent in English.
<sup>2</sup> One-way ANOVAs with Tukey-adjusted comparisons showed the least significant comparison; ns: not significant at p<.05
*Mild acute disease* patients were community-managed with minor, mostly upper respiratory symptoms.
*Severe acute disease*: Patients who had been hospitalized (see <sup>52</sup> for more details)
Medical comorbidities which were investigated/included: chronic cardiac disease (n=10); hypertension (treated) (n=16); obesity (n=7); chronic lung disease (n=1); asthma (treated) (n=11); diabetes (treated) (n=8); chronic kidney disease (n=3); liver disease (n=0); cancer (treated) (n=7); HIV infection (treated) (n=1); transplant recipient (n=1); rheumatologic or autoimmune disorder (n=9); obstructive sleep apnoea (treated but 1, n=3); other comorbidities (n=2: 1 early Parkinson's disease treated)
*Note*: 4 women were pregnant but in the gestation period); 8.7% (n=11) has a history of a psychiatric diagnosis (anxiety disorder and major depressive disorder on treatment)

### Cognitive impairment and overall cognitive performance across time

Detailed cognitive and olfaction data are presented in Table 2. The prevalence of cognitively impaired (practice effect corrected) performance is presented in Figure 1a. In a best fit LME model (Model -2LL=432; AICc=440) with random intercept (p=.003), month post diagnosis was a significant linear predictor of the longitudinal cognitive performance (B=-.025, SE=.004, *p*<.001). Longitudinal cognitive performance declined over time on average by 0.43 [95% CI=-0.39 to -0.47] of a z-score across the post diagnosis period of observation (Figure 1b). In the model where the initial visit cognitive impairment status was added, months post diagnosis (B=-.027, SE=.004, *p*<.001) and impairment status (B=-.768, SE=.092, *p*<.001) were significant predictors of the longitudinal cognitive performance (Model -2LL = 387; AICc = 395). However, the interaction between time and impairment was not significant (*p*=.64). In other words, both impaired and unimpaired longitudinal performance was poorer over time; while those impaired had lower performance overall compared to unimpaired across the entire study period (Figure 1c). Finally, in the best fit longitudinal cognitive model, olfaction, disease severity, lung function, comorbidities, and mental health were not significant as fixed or time interaction predictors.

**Table 2:** Cognition and Olfaction Outcomes by Assessment Time

| Test/measure | 2 Months |  | 4 Months |  | 12 Months |  |
| --- | --- | --- | --- | --- | --- | --- |
|  | *n* | *M (SD)* | *n* | *M (SD)* | *n* | *M (SD)* |
| **Cognition (CogState Computerised Battery)** |  |  |  |  |  |  |
| Detection Speed z-score | 127 | -0.54 (0.70) | 121 | -0.69 (0.65) | 101 | -0.88 (0.76) |
| Identification Speed z-score | 127 | 0.19 (0.59) | 121 | 0.07 (0.64) | 101 | -0.06 (0.62) |
| One Back Speed z-score | 127 | -0.12 (1.13) | 121 | -0.33 (1.01) | 101 | -0.39 (0.98) |
| One Card Learning Accuracy z-score | 127 | 0.63 (1.09) | 121 | 0.09 (1.13) | 101 | 0.10 (1.30) |
| One Back Accuracy z-score | 127 | -0.04 (0.94) | 121 | -0.05 (0.92) | 101 | -0.10 (1.00) |
| Global Mean z-score | 127 | 0.02 (0.50) | 121 | -0.18 (0.52) | 101 | -0.26 (0.55) |
| **Olfaction NIH Odor Identification Test** |  |  |  |  |  |  |
| Global Mean T-score | 126 | 44.20 (10.25) | 119 | 46.05 (9.43) | 93 | 44.00 (9.50) |

Cognition data have been corrected for task-specific practice effects and demographically adjusted for age, sex, and education. Olfaction data is corrected for age, sex, education, and race/ethnicity (i.e., white/other). Two cases had premorbid anosmia and were not tested. At 12 months, 8 cases missed their olfaction assessment due to technical issues.

**Figure 1:**
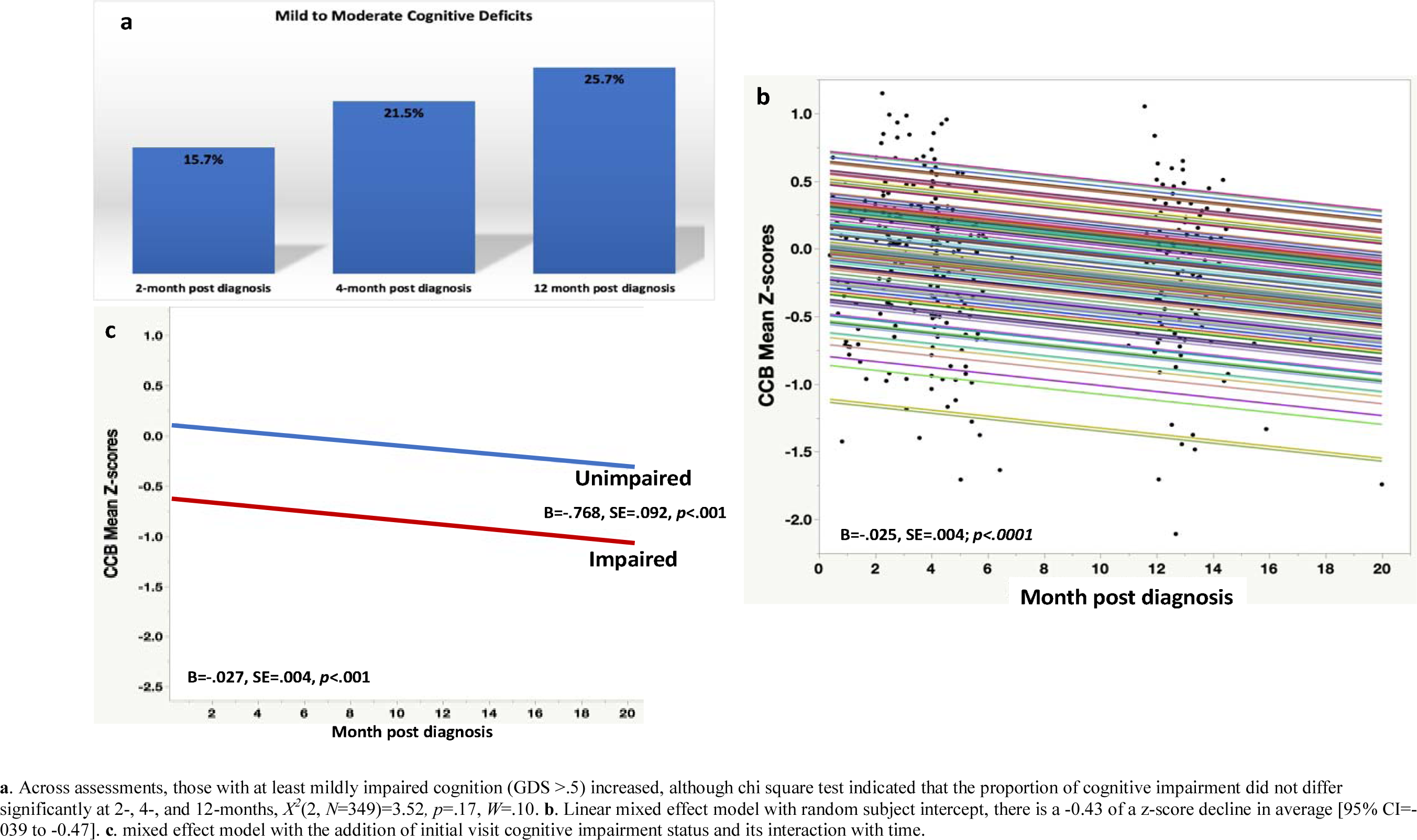
Cognitive impairment and overall cognitive performance over the study period

### Factors associated with mild to moderate cognitive impairment at 2-month post diagnosis

These results are presented in Figure 2. CI was not associated with disease severity (p=.68), lung function (%TLC and %DLCO; p>.30) or having a comorbidity (p=.90). Pre-existing and current mental health were not associated with CI (p=.27 and p=.24 respectively). However, CI was associated with anosmia (*p*=.05) (and at the 2-month post diagnosis visit but not thereafter).

**Figure 2:**
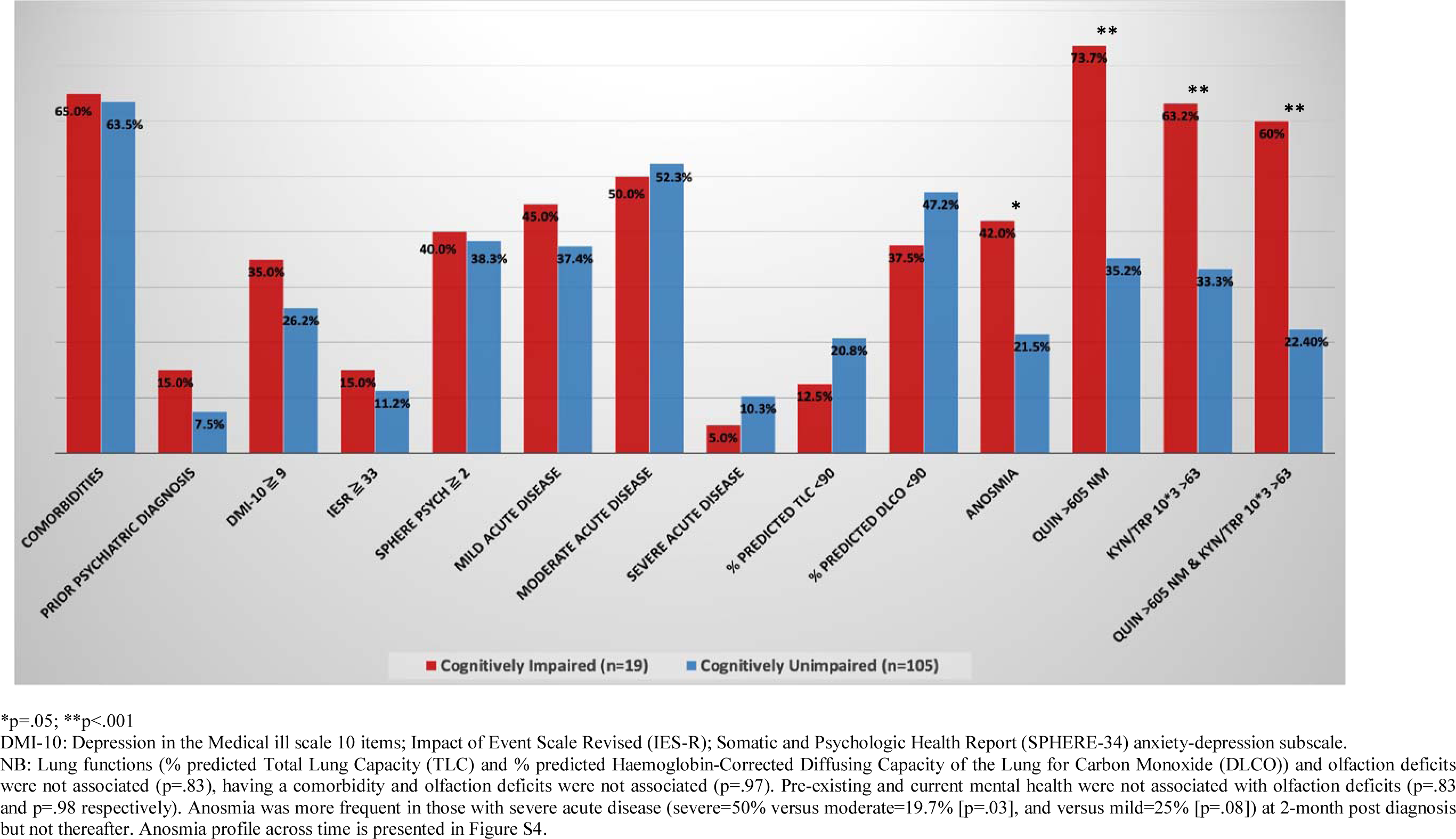
Proportions of medical, clinical symptoms, and clinically relevant KP metabolites’ concentrations in patients with and without Cognitive Impairment 2-month post diagnosis

### Functional status and cognitive impairement

Functional data are provided in Figure S5. Impact of CI on work efficiency and usual activities of daily living was evident at 2- (p=.03) and 4-month post diagnosis (p<.05).

*Within KP dynamics* at 2-month post diagnosis

√KYN, Log 3HK Log QUIN showed moderate to large (r=.42-.75) correlations (Figure S1).

Illustration of the KP dynamics over time is presented in Figure 3. TRP, Log 3HK, and Log PIC showed no significant change over the study period. Both √KYN and KYN as dummy code, (KYN>3 uM=1 versus 0) decreased over time (√KYN: B=-0.086, SE=.007, p<.001; Model -2LL=887, AICc=897; KYN dummy coded: B=-0.47, SE=.005, p<.001; Model - 2LL=565, AICc=574). Log AA decreased over time in a linear pattern at the group level (B=-0.107, SE=.007, p<.001; Model -2LL=998, AICc=1008). Log 3HAA increased over time in a linear pattern at the group level (B=0.24, SE=.007, p<.001; Model -2LL=847, AICc=857). Log QUIN increased in a quadratic pattern both at the group level (significant fixed quadratic time slope, B=-0.13, SE=.002, p<.001), and at the individual level (significant random quadratic time slope p<.001 at each visit) (Model -2LL=498, AICc=512). KP and sex results are provided in the supplementary file.

**Figure 3:**
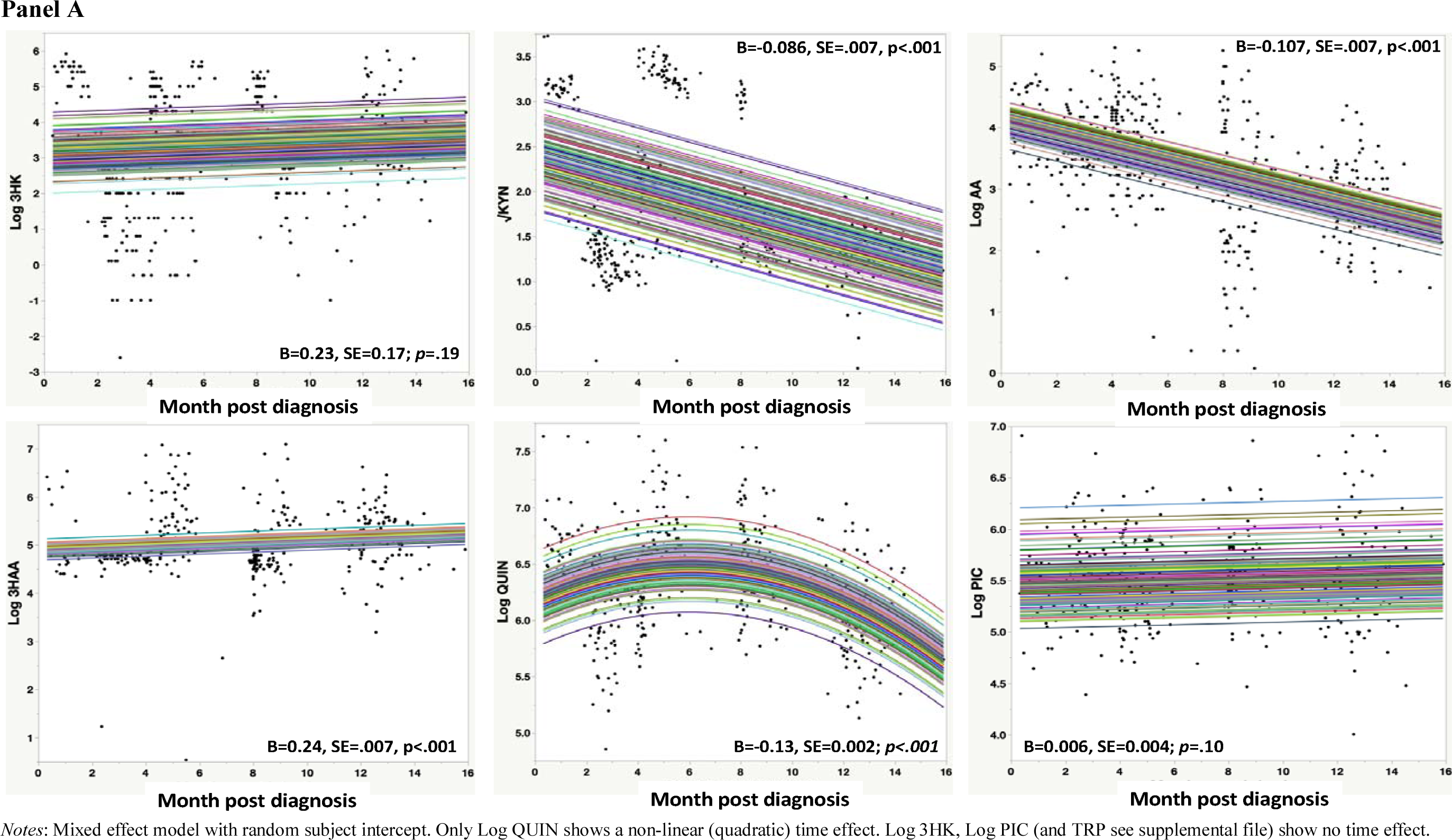
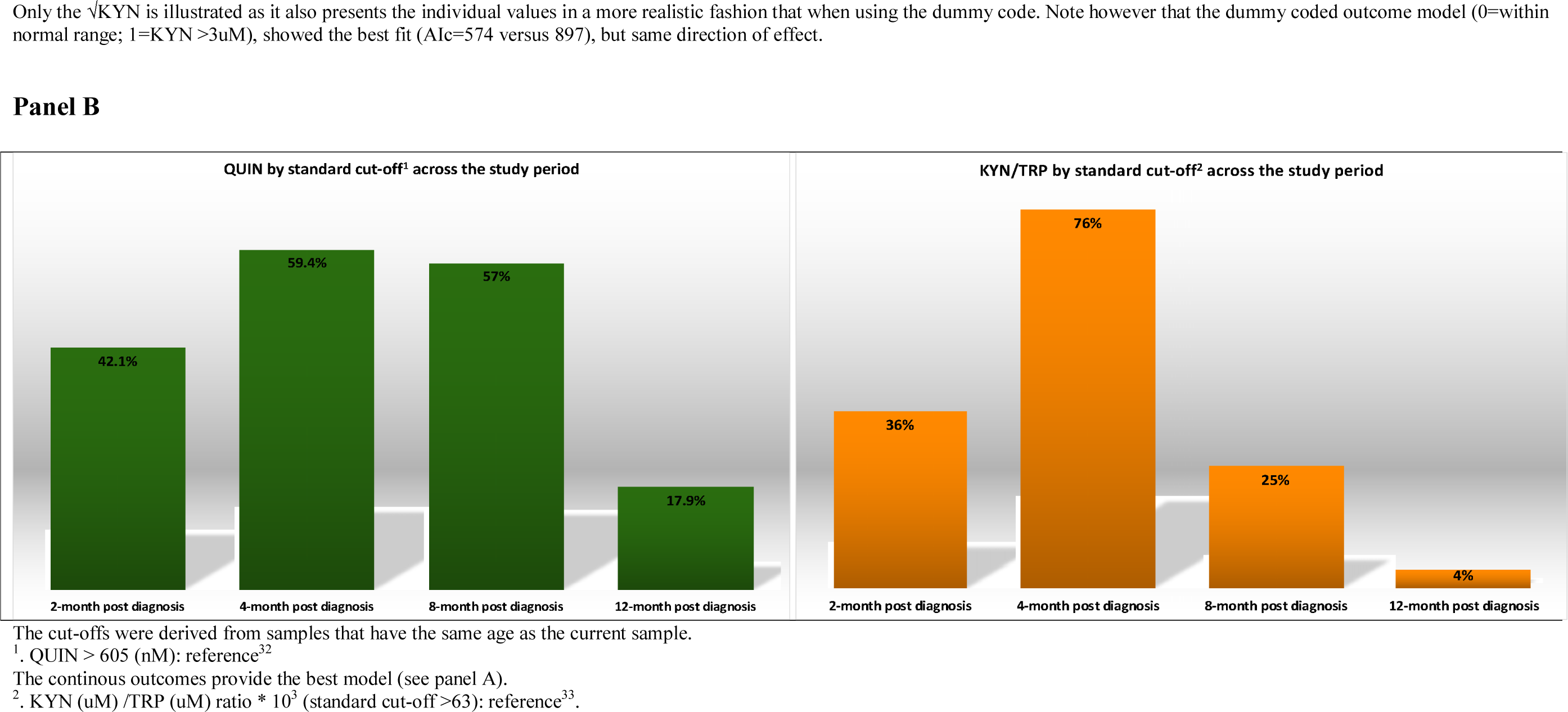
KP dynamics over the study period

### The KP profile and Cognition

Detailed results are presented in Table 4. The best fit model between cognition and the KP were based on a fixed linear time model, with a random intercept. We found a significant negative association between √KYN (FDR significant) or dummy coded KYN, Log 3HK, Log 3HAA (FDR significant), and Log QUIN (FDR significant) and cognition. There were no associations between TRP, Log AA and Log PIC with cognition. Best fit models without the interaction term were not significant, which showed that sex, disease severity, lung function, olfaction, comorbidities, and mental health had no significant associations with cognition over time and did not impact the KP association with cognition.

### QUIN and KYN/TRP ratio and relationship to cognitive impairment at 2-month (time at which the KP was the most activated)

Based on our findings, we explored the ability of elevated QUIN set at a standard cut-off (>605 nM that is 2 SD > age-norms in^39^) and the KYN (uM) /TRP (uM) ratio * 10^3^ (standard cut-off >63^40^) for identifying CI. The advantage of KYN/TRP is that it can be easily obtained in clinical practice. We also tested the combination of having value elevated. These results are part of Figure 2. Overall, cognitively impaired cases have significantly elevated KP metabolites compared to cognitively unimpaired cases at the 2-month post diagnosis visit.

### The KP profile and IFNs

IFN-β was associated with Log QUIN over time in those with persistent lung symptoms (B=- 0.08, SE=.03, p=.005; Model -2LL=321, AICc=335) (Figure S3a). Furthermore, IFN-λ1 showed a trend association with Log QUIN over time also in those with persistent lung symptoms (B=-0.08, SE=.05, p=.05; Model -2LL=323, AICc=338) (Figure S3b). Lastly, IFN-β is associated with √KYN over time in those with persistent lung symptoms (B=-0.07, SE=.03, p=.03; Model -2LL=519, AICc=533) (Figure S3c). Interestingly neither IFN was directly associated with cognition.

The KP profile, Cognition, additional cytokines (IFN-γ, Interleukins, MCP-1, TNF-α), and peripheral brain biomarkers (NFL, S100β, GFAP, GMCSF)

Data for the additional cytokines and peripheral brain biomarkers are provided in Table S2. The analyses showed no associations between any of these biomarkers and cognition except for a negative association at trend level (FDR-corrected) for Log GMCSF and Log MCP-1 (Table S3). Moreover, Log QUIN showed a positive significant association (FDR-corrected, p<.001) with IL-6. Lastly, √KYN showed trend (FDR-corrected) positive associations with Log GMCSF and IL-6 (Table S4).

## Discussion

Our study represents the first comprehensive evaluation of the pathogenesis of post-acute COVID-19 cognitive function and deficits in SARS CoV-2 PCR confirmed unvaccinated patients following predominantly mild to moderate acute COVID-19 disease. The main findings are: 1. Clinically relevant mild to moderate CI affects up to a quarter of patients (Figure 1), and this is associated with lower capacity for returning to pre-COVID work and everyday functioning level up to 4 months post diagnosis (Figure S5); 2. Mild cognitive decline (Figure 1b) is detected independently of whether patients initially showed cognitive impairment, although initially cognitively impaired patients performed worse than cognitively normal patients across the study period (Figure 1c); 3. The KP was activated above the normative age-reference range (Figure 3, panel B). Also, a clear pattern of activation was detected over time with quinolinic acid showing the most dynamic profile (Figure 3 panel A); 4. Mild cognitive decline is uniquely associated with KP activation (Table 3) suggesting a potential causal link thereby indicating it as a biomarker and therapeutic target; 5. Mild cognitive decline was not associated with anxio-depression, olfaction, disease severity, respiratory function, the blood-based biomarkers of brain injury, or any of the cytokines including the interferons.

**Table 3:**
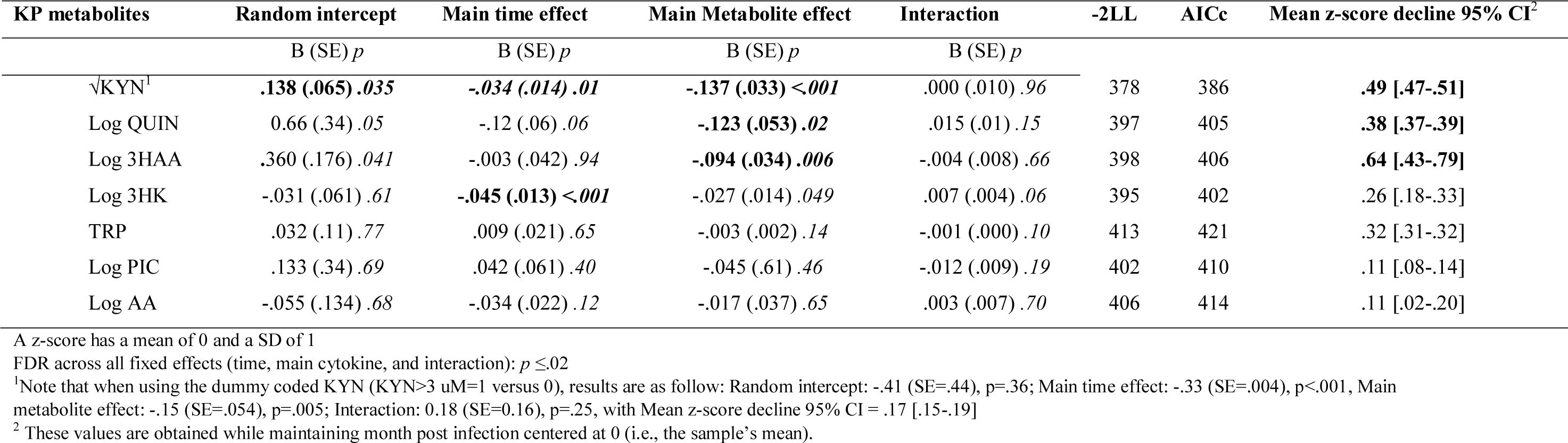
Cognition and KP models organized by effect size (medium to small), confidence of interval on the outcome, and p-value

The potential for the KP to be *causally associated* with mild cognitive decline in the context of this observational study is possible for several reasons^41^. From a statistical perspective^41, 42^, we adjusted for multiple confounding variables which were included as fixed and random effects. Missing data and attrition were small, and all models were adjusted for any residual effects. We measured the major outcomes using standard and robust methodologies. Importantly for cognition, we corrected for the normative effect of age, education, sex, and practice effect mimicking the comparison to a healthy control group. From a conceptual perspective^42^, the mechanism underlying the cause of cognitive decline is plausible according to the current knowledge on the KP and established knowledge on an IFN-mediated pathways, which we demonstrate in this study. The current study was well powered for the continuous data analyses, but not for the next step in testing indices of sensitivity and specificity, and predictive power.

The prevalence of clinically significant CI increased slightly across time (16% to 26%), albeit not to a statistically significant extent. Importantly, the severity of CI was within the mild and moderate deficit range and no patients progressed towards severe cognitive deterioration which is commensurate with what is expected from mild to moderate COVID in the sample’s age range^7, 8^. The detected pattern resembles the “brain fog” that has been widely reported by patients and is now recognized as a long-term consequence of COVID-19 infection^43^. The mild to moderate CI detected via the version of Cogstate battery used involves attention/working memory, psychomotor speed, and new learning. This type of cognitive deficit has been observed in other cognitive studies post mild to moderate COVID-19^7, 8^. Mild to moderate CI can cause difficulties in cognitively demanding tasks and employment^44^, as shown in the current study.

Importantly, there was no association between cognition and anxio-depression, both pre- morbid and current. The relationship between cognition and anxio-depression is complex in viral infection where questions of mental health treatment and recurrence also need to be considered to accurately determine how mental health associates with CI^45^. Objective cognitive performance in COVID-19 and anxio-depression do not have a systematic connection^46^. One explanation for the findings’ discrepancies is probably due to studies’ heterogeneities^47^ in terms of methods and patient’s characteristics (both clinical and socio- economic). Specific studies dedicated to better understanding the connection between mental health and cognition in COVID-19 will be needed. Such studies that will ideally consider a wider socio-economic spectrum than the current study, in addition to pre-morbid mental health status, acute disease severity and ongoing life stressors.

Cognitive performance mildly declined across the 12-months study period by a small to medium effect size decline (-0.43 of a z-score on average). Because the data were corrected for practice effect, in some cases, the patients failed to improve according to expectations based on a practice effect rather declined above and beyond the expected practice effect. Importantly, the detected mild cognitive decline/failure to improve was independent of the initial 2-month visit’s impairment status. Thus, this decline happened across the sample rather than specifically in the cases who had CI at the initial visit. Typically, baseline or initial visit performance is a strong factor in cognitive decline in progressive neurological conditions^37^. Therefore, this finding suggests that an ongoing pathological process affects all patients, and at least partially independently of pre-COVID-19 cognitive functioning. This also means that most impaired patients at the 2-month visit performed in the impaired range across the study timeline representing persistent CI. This contrasts against studies which have focused on severe cases of acute COVID-19 where pre-morbid neurological conditions predict cognitive deterioration^3^. Our finding of cognitive decline/failure to improve is in accordance with emerging cognitive^3, 48^ and brain imaging^49^ studies assessing post-acute mild-moderate COVID-19 patients at 12-months post diagnosis. Studies with longer follow-up will be critical to determine if this trend is maintained or whether patients eventually improve^50^. This will be particularly important in older participants with acute mild disease entering the dementia age range who, according to a large Chinese study^48^, are at risk of early onset cognitive decline. The use of optimal methods to detect cognitive decline will be paramount including the use of tests specifically designed to measure cognitive changes rather than instruments designed for cross-sectional assessment with restricted range of values such as the MoCA or MMSE^16^.

Previous cognitive studies on COVID-19-associated impairment rate are difficult to compare with our study. Many had CI rates that were substantially higher (Table S1) as they included predominantly hospitalized patients with higher rates of comorbidities than seen in our patients. Additionally, we corrected for the normative effects of age, education, and sex unlike most studies. Becker *et al.*^51^, who corrected for demographics in their neuropsychological tests found impairment rates more consistent with the current study: 13 to 39% of hospitalized patients were overall impaired, 6 to 26% of those presenting to emergency department, and 5 to 16% in those managed in the community.

In the current study, olfaction and cognitive change had different dynamics, but were initially associated at the 2-month post diagnosis visit. The pathogenetic significance of this is unclear but the lack of association on subsequent visits argues against an olfactory nerve brain entry route in accordance with other literature^52^.

Moreover, whilst disease severity was associated with reduced lung function, there was no association between disease severity or lung function with mild cognitive decline, suggesting that symptoms of acute COVID-19 infection are not necessarily associated with cognitive function in the months following COVID-19 infection. Our finding of no association between CI and disease severity including respiratory function is in keeping with other studies (Table S1)^2^. This finding has major public health and economic implications: relying on disease severity (especially mild to moderate distinction) to estimate disease burden will result in a significant underestimate.

The KP is upregulated in this cohort of patients recovering from mild to moderate COVID- 19, most significantly in relation to mild cognitive decline. Indeed, KP metabolites (3HAA, QUIN and KYN) were the only blood markers associated with mild cognitive decline. While other studies have observed KP upregulation during acute infection and thereafter, none has explored the relationship to cognitive function. Moreover, KP activation was associated with IFN-β, a known inducer of the KP reinforcing the biological plausibility of our findings^24^. Normative reference range for the KP are known^39, 40^ and we show that up to 60% of the sample have an activated KP using QUIN age-norms standards (Figure S3). QUIN was the most dynamic KP metabolite with a curvilinear time pattern suggesting its peak concentration was captured. Other metabolites also significantly changed over time further demonstrating an overall upregulation of the KP. KYN and 3HAA significantly decreased across time without a curvilinear profile, possibly because their peak concentrations were earlier or because there have truly have a linear trajectory. The dynamism of the KP makes it a robust maker longitudinally. Cross-sectionally at 2-month post diagnosis, we can nevertheless detect the KP association with CI for QUIN and the easily measured KYN/TRP^40^ when normative reference range are considered. KYN/TRP could be useful in clinical practice to detect the most clinically impaired cases.

Critically, no other tested variables were associated with cognitive decline including lung function, longitudinal olfaction performance, and mental health. Further, none of the tested Interleukins, inflammatory biomarkers, and peripheral biomarkers of brain injury were associated with cognitive decline. This contrasts with other studies that have noted abnormalities in these biomarkers with acute infection and severe disease^28–31, 53^.

KP activation is biologically and pathogenetically significant as it is known to be monocyte-macrophage mediated, leading to immune tolerance and neurotoxicity if persistently activated^22^. Further, recent evidence points to KP activation having antiviral effects through QUIN induced production of type I interferon via N-methyl-d-aspartate receptor activation with Ca2+ influx activating Calcium/calmodulin-dependent protein kinase /interferon regulatory factor 3^54^. Indeed, our results are in keeping with the recent finding that the monocyte-macrophage is the primary driver in COVID-19 pathogenesis^55^ as this is the only cell type with complete KP expression and capability of QUIN production^22^. KP-associated CI have been detected in other viral infections such as HIV, influenza, SARS, and Middle East respiratory syndrome^1, 22^. Whilst we did not examine the CSF in our patients it is well established that some KP products can cross an intact blood brain barrier and then serve as extra substrates for QUIN production by perivascular macrophages and microglia^22^. Moreover, there is some evidence that chronic exposure to QUIN can damage the blood brain barrier allowing ingress of immune related toxins including KP products and QUIN^56^ traffic to the CNS where QUIN has a neurotoxic effect. KP modulating drugs are in various stages of development but hold promise for clinical use^57^.

Our study has several limitations. Our sample size was smaller in comparisons to some large studies (N>500) that have been published, but this is tempered by a longitudinal design with at least three visits (total cognitive and KP datapoints=349-512) where statistical power is drawn from repeated testing. Further, the study includes a small sub-sample for the severe cases, and it is possible that some analyses were under-powered. However, the previous literature has concentrated on severe cases, and it is obvious from this research that severe COVID pathogenesis differs from that of mild-moderate cases particularly in those with high comorbidity burden. Our sample is restricted to the socio-economically advantaged parts of Sydney and with relatively high education (15 years on average), optimal care access and management of medical comorbidities^32^. Therefore, generalization of the current findings to less advantaged populations is not possible. Nevertheless, if we consider this population as a “healthy” benchmark, we expect worse cognitive outcomes in more comorbid and more vulnerable populations in agreement with the existing literature. We do not yet have longitudinal data for mental health, and lung function, but close to acute effects regarding these conditions are the strongest according to previous findings^20, 58^ and worsening of pre-morbid anxio-depression during the pandemic is not systematic^58^ but is associated with social support and living conditions^59^. We had restricted data for IFN-β and IFNλ1, however the connections between the KP and IFNs are well established^24^. We used a relatively short cognitive battery, but it targeted cognitive functions that are primarily impaired in patients recovering from COVID-19^16^. The study has no contemporaneous healthy controls because of the infectious challenges of the pandemic. However, we used normative data for cognition that was closely comparable to the demographics of the current sample, including for normative correction practice effect. Further, the immunological findings underpinning IFNs were compared to demographically matched controls with other types of coronaviruses^20^. Lastly, we provided several lines of evidence for the upregulation of the KP (Figure S2). The cohort was unvaccinated at the time of the assessment and further assessment are planned post-vaccination. Our results remain potentially relevant to vaccinated patients who still develop mild and moderate acute COVID-19 disease, although assessment of cognition and the KP are warranted in these patients.

In conclusion, KP activation was found to be the first biomarker for post-acute COVID-19 cognitive impairment/decline, highlighting the KP as a monitoring tool and therapeutic target.

## Data Availability

All data produced in the present study are available upon reasonable request to the authors

## Acknowledgements

We thank the participants for their time.

Thanks to the ADAPT nursing team and operation team for outstanding services.

