## Supplemental files for "Post-acute COVID-19 cognitive impairment and decline uniquely associate with kynurenine pathway activation: a longitudinal observational study"

**Table S1: Summary of Studies Objectively Assessing Cognitive Functioning Post-Acutely in COVID-19 Patients**

| Study | Sample | Design | Timeline | Measures | Main Findings |
|---|---|---|---|---|---|
| **Alemanno et al., 2021** Italy | 87 patients admitted to the COVID-19 Rehabilitation 4 groups according to the respiratory assistance in the acute phase: Group1 (orotracheal intubation), Group2 (non-invasive ventilation using Biphasic Positive Airway Pressure), Group3 (Venturi Masks), Group4 (no oxygen therapy) 62 Male, mean age 67.23 ± 12.89 years | Longitudin al | Follow-ups were performed at one month after home-discharge 56 patients (22 of Group 1, 12 of Group 2, 20 of Group 3 and 2 of Group 4) | MMSE, MoCA, Hamilton Rating Scale for Depression, and Functional Independence Measure (FIM) | MoCA scores showed that 74.2% Group 1, 94.4% Group 2, 89.6% Group 3 and 77.8% presented with deficits MMSE 12.9% Group 1 had mild to severe deficits; 55.6% Group 2 had mild to moderate deficits; 48.3% Group 3 had mild to severe deficits; and 44.4% had moderate deficits. Results were correlated with age At follow-up overall improvement on MoCA and MMSE, but many deficits remained. Group 1 is the least impaired. |
| **Almeria et al. (2020)** Spain | 35 hospitalised patients, 54% female. 20% in ICU, 60% oxygen treatment, 20% neither ICU or oxygen treatment. Mean age 47.6 years (SD = 8.9 years) exclusion of subjects older than 60 or with psychiatric condition | Cross-sectional | 10–35 days post hospital discharge | Visual Reproduction (Wechsler Memory Scale – IV), Verbal Learning via list learning, interference, and recognition | Patients requiring oxygen therapy during hospitalization: impairment in attention, working memory, processing speed, executive function, and global cognition; ICU treated patients: impaired executive dysfunction |
| **Amalakanti et al. (2021)** India | 93 asymptomatic COVID-19 patients, 52.3% female, mean age 36.2 years (SD = 11.7 years). 102 healthy controls, 54.7% female, mean age 35.6 (SD = 9.8 years). All patients had less than four years of education | Cross-sectional | Unclear if measured post-acutely as details not provided | MoCA | Differences in overall MoCA scores between groups was negligible, although COVID-19 performed significantly lower in visuoperception, naming and fluency domains than that of controls. |
| **Beaud et al. (2021)** Switzerland | 13 previous ICU patients in in-patient rehabilitation, 23% female. Mean age 64.8 years (SD = 7.6 years) patients still in hospital. Exclusion of patients with pre-existing psychiatric diseases, not controlling for age in FAB | Cross-sectional | Post-critical acute stage of severe COVID-19 (still in hospital) | MoCA, Frontal Assessment Battery (FAB) | Cognitive impairment in 69% (31% mild, 39% moderate to severe). More extensive impairment in executive, memory, attentional and visuospatial functions. 92% impaired in lexical fluency (FAB subtest) |
| **Becker et al. (2021)** United States | 740 patients, 51% outpatients, 22% emergency department, 27% hospitalised. 63% female, | Cross-sectional | 7.6 months (SD = 2.7) after SARS-CoV-2 diagnosis | Number Span Forward and backward, Trail Making Test Part A and B, | Deficits across domains: processing speed (18%), executive functioning (16%), phonemic fluency (15%), |

**Table S1: Summary of Studies Objectively Assessing Cognitive Functioning Post-Acutely in COVID-19 Patients**

| Study | Sample | Design | Timeline | Measures | Main Findings |
|---|---|---|---|---|---|
| | Mean age 49 years (SD = 14.2 years), education: 14% ≤12 years education, 86% > 12 years education | | | Phonemic and Category Fluency, Hopkins Verbal Learning Test-Revised. Did not account for mental health | category fluency (20%), memory encoding (24%), and memory recall (23%). Compared to outpatients, hospitalised patients were more likely to have attention, executive function, category fluency and memory impairments |
| **Bungenberg et al. 2021** Germany | 50 patients with persisting symptoms for at least 4 weeks were included and classified by initial hospitalization status: hospitalized (*n* = 21) and non-hospitalized (*n* = 29). Median Age 50.5 years (range 22–84 years), 56% female; median education: 15.5 (12.75–18) | Cross-sectional | Median time from SARS-CoV-2 detection to investigation was 29.3 weeks (range 3.3-57.9) | MoCA, version 7 Test of Attentional Performance (TAP), TMT A and B, Digit span forwards and backwards. Verbal fluencies, Stroop test variant (Farbe-Wort-Interferenztest, FWIT), Auditory Verbal Memory Test (VLMT), Rey Complex Figure copy and recall, Boston Naming Test | Mild deficits were found in attention, executive functions, and memory Hospitalized patients performed worse in global cognition, logical reasoning, and processes of verbal memory. In both groups, fatigue severity was associated with reduced performance in attention and psychomotor speed tasks and reduced quality of life and with more persisting symptoms. |
| **De Lorenzo et al. (2020)** Italy | 185 patients, 34% female. 68% hospitalised, 32% discharged from emergency department. Mean age 57 years (SD = 10.4 years) Only patients with suspected pneumonia | Cross-sectional | 3.3 weeks post-discharge from hospital or emergency department | MoCA Did not account for mental health | Cognitive impairment in 25% |
| **Del Brutto et al. (2021 & 2022)** Ecuador | 2021: 52 non-hospitalised patients, 63% female. Mean age 62.6 years (SD=11 years). 41 healthy, demographically matched controls. Including non-hospitalised people aged 40+ years | Longitudinal | First assessment: 2013-2015, Second Assessment: 2017-2019, Third Assessment: 6 months post-infection | MoCA – Spanish Version (Adjusted for demographics, cardiovascular risk factors, mental health, and education) | COVID-19 patients 18 times more likely to develop cognitive decline compared to healthy controls. 12% of patients declined cognitively after infection compared to 2% of healthy controls assessed on same timeline. |
| | 2022: 78 participants, 50 with history of mild COVID-19 and 28 without | Longitudinal | 2 cognitive testing before the pandemic and 2, 6 and 18 months after the initial SARS-CoV-2 outbreak | Same | Significant-likely age-related-decline in MoCA scores between the two prepandemic tests which did not differ between groups; at 6 months, only COVID-19 survivors showed significant decline, but reversed at 18-month |
| **Frontera et al. (2022)** USA | N=242 patients (median age 65, 64% male, 34% intubated during hospitalization) and | Longitudinal | 6- to 12-months post infection | Modified Rankin Score, T-MoCA, Neuro-QoL | Those with neurological complications (N=113) had higher fatigue scores but other differences. |

**Table S1: Summary of Studies Objectively Assessing Cognitive Functioning Post-Acutely in COVID-19 Patients**

| Study | Sample | Design | Timeline | Measures | Main Findings |
|---|---|---|---|---|---|
| | N=174 completed both 6- and 12-month follow-up | | | Results adjusted for age, sex, race, pre-COVID Modified Ranking Score and intubation status | Significant improvements in outcome trajectories from 6- to 12-months were observed in T-MoCA scores (56% improved, median difference 1 point. and Neuro-QoL anxiety scores (45% improved). Non-significant improvements occurred in fatigue, sleep and depression. |
| **Hampshire et al (2021)** UK | N= 81,337 | Cross-sectional | January and December 2020 | Clinically validated web-optimized assessment (gbit.cognitron.co.uk), and questionnaire items capturing self-report of suspected and confirmed COVID-19 infection and respiratory symptoms | COVID+ showed significant cognitive deficits versus controls when controlling for age, gender, education level, income, racial-ethnic group, pre-existing medical disorders, tiredness, depression and anxiety, and pre-morbid level. Hospitalized worse than non-hospitalized and controls; non-hospitalized worse than controls |
| **Hellgren et al. (2021).** Sweden | 35 hospitalised patients, 20% female. Median age 59 years (IQR: 51-66), 19 needed mechanical ventilation, 14 did not require ventilation. Only including those with concerning results on neuropsychological testing or suspected cognitive impairment | Cross-sectional | Neurocognitive testing: 5 months post-discharge (median 142 days); MRI: 7 months post-discharge (median 217 days). | Repeatable battery for the assessment of neuropsychological status (RBANS) | Cognitive impairment in 46%, of which 29% had severe impairment. Patients with abnormalities on MRI had a significantly (p = .031) lower visuospatial index compared to those with a normal MRI. |
| **Jaywant et al. (2021)** United States | 57 previous ICU patients in in-patient rehabilitation, 25% female. Mean age 64.5 years (SD = 13.9 years) Only including patient with suspected cognitive impairment | Cross-sectional | 2 months post hospital admission, still in in-patient rehabilitation | Age-normed Brief Memory and Executive Test (BMET) | Cognitive impairment in 81%, ranging from mild to severe. Common deficits in working memory (55%), set-shifting (47%), divided attention (46%), and processing speed (40%) |
| **Liu et al., 2022** China | 1438 COVID-19 survivors and 438 control individuals; *all aged 60+*; COVID-19 was categorized as severe or nonsevere following the American Thoracic Society guidelines | Longitudinal | Follow-up at 6 and 12 months | Informant Questionnaire on Cognitive Decline in the Elderly and the Telephone Interview of Cognitive Status-40 | The incidence of cognitive impairment in survivors 12 months after discharge was 12.45%. Individuals with severe cases had lower Telephone Interview of Cognitive Status-40 scores than those with nonsevere cases and control individuals at 12 months. Severe COVID-19 was associated with a higher risk of early-onset cognitive decline, late-onset |

**Table S1: Summary of Studies Objectively Assessing Cognitive Functioning Post-Acutely in COVID-19 Patients**

| Study | Sample | Design | Timeline | Measures | Main Findings |
|---|---|---|---|---|---|
| | | | | | cognitive decline and progressive cognitive decline while nonsevere COVID-19 was associated with a higher risk of early-onset cognitive decline (adjusted for age, sex, education level, body mass index, and comorbidities) |
| **Mattioli et al. (2021)** Italy | 120 health care workers who had COVID-19 of mostly mild to moderate severity (2 patients required hospitalisation), 75% female, mean age 47.9 years, mean education 16 years. 30 (controls) health care workers who had not had COVID-19, 73.3% female, mean age 45.7 years, mean education 18 years. | Cross-sectional | Mean of 126 days from diagnosis (range of 12 to 215 days) | Controlled Oral Word Association (COWA), Rey figure copy and recall, California Verbal Learning Test (CVLT), TEA attention test, Tower of London test, MMSE, Depression anxiety and stress scale-21 (DASS-21) | Frequency of neurological deficits and cognitive impairment was negligible. COVID-19 patients were impaired on 1.6 tests on average, and this was not significantly different to those who did not have COVID-19. Anxiety, stress and depression was significantly more elevated in those with COVID-19 than those without COVID-19. |
| **Mattioli et al. (2022)** Italy | 52 COVID+ cases treated in intensive care unit (ICU patients), 163 not hospitalized (non-ICU patients). | Cross-sectional | 4 months after the diagnosis | Neurological exam and extensive cognitive evaluation, investigating general cognition, memory, verbal fluency, visuospatial abilities and executive functions | eripheral nervous system deficits in 2/163 (1.2%) of non-ICU and in 7/52 (13.5%) of the ICU cases; ICU patients performed significantly worse than non-ICU cases; unrelated to tested comorbid effects |
| **Mazza et al. (2021)** Italy | 226 patients who initially presented to emergency department, 177 hospitalised, 49 discharged and treated at home. 34% female, mean age 58.5 years (SD = 12.8 years). Only subsample of 130 patients completed BACS.<br>Exclusion of patients older than 70 | Longitudin al | 1 and 3 months after hospital discharge | Brief Assessment of Cognition in Schizophrenia (BACS) – age normed<br>Demographics not provided for sample who completed BACS. | 78% of sample had poor performances in at least one cognitive domain, with executive functions and psychomotor coordination impaired in 50% and 57% of the sample respectively at 3 month follow up. Self-reported psychopathology influenced cognition. |
| **Miskowiak et al. (2021)** Denmark | 29 hospitalised patients, 41% female. 100 healthy age- and education-matched controls | Longitudin al (baseline results) | 3-4 months post hospital discharge | Screen for Cognitive Impairment in Psychiatry - Danish Version (SCIP-D), and the Trail Making Test-Part B.<br>Did not control for mental health | Cognitive impairment in 59%- 65% (depending on which cutoff was applied). Large effect size for verbal learning and executive functioning. Moderate impairment for working memory, verbal fluency and psychomotor speed |

**Table S1: Summary of Studies Objectively Assessing Cognitive Functioning Post-Acutely in COVID-19 Patients**

| Study | Sample | Design | Timeline | Measures | Main Findings |
|---|---|---|---|---|---|
| **Raman et al. (2021)** UK | 58 hospitalised patients, 41% female. 95% mechanical ventilation, 36% ICU. Mean age 55.4 years (SD = 13.2 years). 30 uninfected controls, matched for age, sex, BMI and comorbidities | Longitudinal (baseline results) | 2.3 months from disease-onset, (1.6 months post hospital discharge) | MoCA | Impairments in executive and visuospatial domains significantly more common in patients than controls (40% patients vs 16% controls), median MoCA scores in patients not significantly different from controls |
| **Soldati et al. (2021)** Brazil | 23 ICU patients, 22% female. Median age 53.6 years (IQR ± 11.7)<br>Only ICU patients with mechanical ventilation | Cross-sectional | 3.3 months post hospital discharge | Telephone Interview of Cognitive Status (TICS)<br>Did not control for mental health and education | 13% met criteria for mild cognitive impairment, 61% of patients fell within normal limits on cognitive assessments, 26% had ambiguous result. No participants had severe cognitive dysfunction |
| **Tomasoni et al. (2021)** Italy | 25 hospitalised patients with virological clearance<br>Time of hospitalisation, disease severity and demographic information not specified | Cross-sectional | 1 – 3 months after virological clearance | MMSE (adjusted for age and education years)<br>Did not control for mental health | Cognitive impairment in 40%, ranging from mild to severe. 16% had ambiguous results |
| **Woo et al. (2020)** Germany | 18 patients, 58% female. 61% hospitalised, 39% non-hospitalised, no ICU. Mean age 42.2 years (SD=14.3 years). 10 age-matched control. Controls not matched for sex | Cross-sectional | 2.8 months post recovery | Modified Telephone Interview of Cognitive Status (TICS-M)<br>Did not account for control for education. | Patients scored significantly lower on TICS-M compared to healthy controls, especially regarding short-term memory, attention, concentration and language. Neuropsychologic deficits were independent from hospitalization and disease severity. |
| **Zhou et al. (2020)** China | 29 patients, 38% female. Median age 47 years (SD = 10.54 years). 29 age-, sex- and education-matched controls.<br>Disease severity not reported<br>Only patients with >9 years education.<br>Excluded patient with a mental health disorder and left handedness | Cross-sectional | 2-3 weeks post infection | iPad-based online neuropsychological tests, including the Trail Making Test (TMT), Sign Coding Test (SCT), Continuous Performance Test (CPT), and Digital Span Test (DST). | Mild cognitive impairments among patients, mainly affecting sustained attention. No significant difference between patient and healthy controls in TMT, SCT, or DST |

*Note.* This tables features studies assessing adult patients with confirmed SARS-CoV-2 infection (according to World Health Organization (2021a) criteria), using objective cognitive assessments, censored 31st of March 2022. Methodology of this literature review included searching on PubMed Database and Google Scholar using the following keywords: neurocognitive functioning COVID-19, cognitive functioning COVID-19, SARS-CoV-2 cognition, and COVID-19 cognition. Included in literature review were papers with adult participants who had a confirmed diagnosis of COVID-19, journals written in English, and studies which used standardised and objective measures. Self-report measures were not included. IQR = Interquartile Range, SD = standard deviation, Montreal Cognitive Assessment (MoCA), Mini-Mental State Exam (MMSE).

**Table S2: KP metabolites and cytokine log-transformed values over time.**

| Biomarker | 2-month post diagnosis | 4-month post diagnosis | 8-month post diagnosis | 12-month post diagnosis | Lower limit of detection |
|---|---|---|---|---|---|
| n | 126 | 126 | 114 | 84 | |
| | M (SD) | M (SD) | M (SD) | M (SD) | |
| **KP** | | | | | |
| TRP | 49.44 (10.43) | 48.61 (10.53) | 47.71 (7.06) | 48.09 (11.33) | |
| √KYN | 1.83 (0.86) | 2.41 (0.82) | 1.69 (0.58) | 1.28 (0.29) | |
| Log 3HK | 2.63 (1.86) | 3.70 (1.62) | 3.46 (1.45) | 3.55 (1.14) | |
| Log AA | 3.76 (0.58) | 3.69 (0.85) | 3.11 (1.10) | 2.72 (0.59) | |
| Log 3HAA | 4.89 (0.52) | 5.09 (0.75) | 5.02 (0.60) | 5.17 (0.61) | |
| Log QUIN | 6.30 (0.50) | 6.49 (0.40) | 6.46 (0.43) | 6.03 (0.40) | |
| Log PIC | 5.51 (0.38) | 5.48 (0.38) | 5.51 (0.34) | 5.60 (0.56) | |
| **Cytokines** | | | | | |
| Log NFL | 1.95 (0.74) | 1.96 (0.77) | 2.27 (0.73) | 2.82 (1.20) | -1.93 |
| Log GFAP | 6.30 (1.77) | 6.21 (1.67) | 6.36 (1.86) | 6.15 (2.04) | 0.48 |
| Log S100B | 3.02 (0.57) | 3.05 (0.78) | 3.17 (0.84) | 3.13 (1.46) | -0.08 |
| Log GMCSF | -0.79 (3.44) | -0.13 (3.58) | 0.03 (3.50) | -0.54 (3.70) | -4.61 |
| Log IFNγ | -0.10 (1.20) | -0.13 (1.34) | -0.15 (1.14) | 0.03 (1.16) | -3.00 |
| Log IL1β | 3.16 (1.31) | 3.08 (1.43) | 3.17 (1.37) | 3.32 (1.25) | 0.00 |
| Log IL1Ra | 2.03 (0.98) | 1.79 (1.10) | 1.82 (1.03) | 2.01 (0.92) | -0.69 |
| Log IL2 | -0.75 (1.86) | -0.88 (2.07) | -0.60 (1.90) | -0.41 (1.88) | -3.51 |
| Log IL4 | -2.43 (2.08) | -2.39 (2.17) | -2.57 (1.97) | -2.46 (2.14) | -3.91 |
| Log IL5 | 1.05 (0.79) | 0.86 (0.86) | 0.94 (0.75) | 0.97 (0.88) | -1.31 |
| Log IL6 | -0.42 (1.22) | -0.48 (1.31) | -0.46 (1.19) | -0.42 (1.00) | -3.91 |
| Log IL8 | 2.00 (0.72) | 1.82 (0.75) | 1.85 (0.79) | 1.83 (0.79) | -1.90 |
| Log IL10 | -1.32 (2.30) | -1.81 (2.26) | -1.70 (2.31) | -1.32 (2.36) | -3.91 |
| Log IL12p40 | 4.44 (0.77) | 4.25 (0.83) | 4.23 (0.81) | 4.48 (0.85) | 1.85 |
| Log IL12p70 | -0.28 (2.00) | -0.34 (2.09) | -0.29 (1.83) | -0.17 (1.89) | -2.30 |
| Log IL13 | 2.40 (2.02) | 2.10 (2.26) | 2.16 (2.10) | 2.26 (2.16) | -1.14 |
| Log TNFα | 3.39 (0.71) | 3.25 (0.74) | 3.29 (0.62) | 3.41 (0.60) | 0.27 |
| MCP-1 | 268.50 (102.56) | 242.46 (87.66) | 254.33 (93.76) | 246.13 (89.82) | 54.46 |
| IFN-β | 3.84 (1.15) * | | | | |
| IFN-λ1 | 3.00 (0.85) * | | | | |

KP values did not have a lower limit of detection. * n = 57, only measured at time 1.

**Table S3. Effect on Cognition by biomarker and biomarker * time interaction**

| Biomarker | Random intercept | Main time effect | Main Biomarker effect | Interaction | -2LL | AICc |
|---|---|---|---|---|---|---|
| | B (SE) p | B (SE) p | B (SE) p | B (SE) p | | |
| Log NFL | **.173 (.028) <.001** | -.034 (.013) .007 | -.008 (.029) .773 | .005 (.005) .355 | 450 | 460 |
| Log GFAP | **.14 (.029) <.001** | -.021 (.019) .283 | -.020 (.022) .362 | -.001 (.003) .766 | 221 | 231 |
| Log S100B | **.174 (.028) <.001** | -.025 (.016) .112 | .022 (.032) .485 | .001 (.005) .920 | 450 | 460 |
| Log GMCSF | **.172 (.028) <.001** | **-.023 (.004) <.001** | -.020 (.009) *.024* | -.001 (.001) .592 | 410 | 420 |
| Log IFNγ | **.175 (.028) <.001** | -.023 (.004) <.001 | -.033 (.026) .198 | -.006 (.003) .112 | 407 | 417 |
| Log IL1β | **.172 (.028) <.001** | -.013 (.012) .271 | -.015 (.026) .574 | -.003 (.003) .366 | 410 | 420 |
| Log IL1Ra | **.172 (.028) <.001** | -.017 (.010) .080 | .018 (.032) .583 | -.003 (.005) .515 | 409 | 419 |
| Log IL2 | **.173 (.028) <.001** | **-.024 (.004) <.001** | -.015 (.017) .373 | -.003 (.002) .244 | 411 | 421 |
| Log IL4 | **.173 (.028) <.001** | **-.024 (.006) <.001** | .001 (.017) .938 | <.001 (.002) .943 | 413 | 423 |
| Log IL5 | **.171 (.028) <.001** | **-.024 (.006) <.001** | -.011 (.037) .775 | .001 (.005) .806 | 409 | 419 |
| Log IL6 | **.175 (.028) <.001** | **-.025 (.004) <.001** | -.043 (.028) .132 | -.005 (.004) .221 | 408 | 418 |
| Log IL8 | **.172 (.028) <.001** | -.019 (.011) .095 | .015 (.045) .732 | -.002 (.006) .713 | 409 | 419 |
| Log IL10 | **.172 (.028) <.001** | **-.028 (.005) <.001** | <.001 (.015) .997 | -.003 (.002) .055 | 410 | 420 |
| Log IL12p40 | **.17 (.028) <.001** | -.018 (.023) .417 | .071 (.042) .088 | -.001 (.005) .815 | 407 | 417 |
| Log IL12p70 | **.171 (.028) <.001** | **-.023 (.004) <.001** | -.027 (.016) .103 | <.001 (.002) .943 | 410 | 420 |
| Log IL13 | **.172 (.028) <.001** | **-.020 (.006) .001** | -.003 (.015) .859 | -.001 (.002) .505 | 413 | 423 |
| Log TNFα | **.173 (.028) <.001** | -.017 (.022) .458 | -.005 (.049) .923 | -.002 (.007) .770 | 409 | 419 |
| MCP-1 | **.173 (.028) <.001** | -.026 (.012) .028 | .001 (<.001) .047 | <.001 (<.001) .679 | 425 | 435 |

FDR across all fixed effects (time, main cytokine, and interaction): $p<0.001$

#### Table S4. Effect on <u>KP metabolites (KYN and QUIN)</u> by biomarker and biomarker * time interaction

| KP metabolite | Biomarker | Random intercept | Main time effect | Main biomarker effect | Interaction | -2LL | AICc |
|---|---|---|---|---|---|---|---|
| | | B (SE) p | B (SE) p | B (SE) p | B (SE) p | | |
| **√KYN** | Log NFL | - | **-.077 (.018) <.001** | .032 (.044) .474 | -.004 (.008) .584 | 900 | 910 |
| | Log GFAP | - | **-.113 (.03) <.001** | .015 (.025) .546 | .003 (.005) .47 | 529 | 539 |
| | LogS100B | - | **-.068 (.025) .006** | .007 (.043) .878 | -.006 (.008) .449 | 899 | 909 |
| | Log GMCSF | - | **-.085 (.007) <.001** | .031 (.009) .001 | -.003 (.002) .107 | 892 | 902 |
| | Log IFNγ | - | **-.086 (.007) <.001** | .008 (.026) .754 | .007 (.005) .171 | 896 | 906 |
| | Log IL1β | - | **-.109 (.018) <.001** | .008 (.024) .741 | .007 (.005) .173 | 897 | 907 |
| | Log IL1Ra | - | **-.117 (.015) <.001** | -.024 (.032) .461 | .016 (.007) .021 | 893 | 903 |
| | Log IL2 | - | **-.085 (.007) <.001** | .009 (.017) .608 | .001 (.004) .733 | 901 | 911 |
| | Log IL4 | - | **-.081 (.01) <.001** | -.001 (.016) .963 | .002 (.003) .569 | 902 | 912 |
| | Log IL5 | - | **-.101 (.01) <.001** | -.031 (.039) .427 | .016 (.008) .049 | 894 | 904 |
| | Log IL6 | - | **-.086 (.007) <.001** | .070 (.027) .009 | -.003 (.006) .568 | 892 | 902 |
| | Log IL8 | - | **-.103 (.018) <.001** | -.037 (.043) .391 | .009 (.009) .321 | 897 | 907 |
| | Log IL10 | - | **-.081 (.008) <.001** | .009 (.014) .540 | .004 (.003) .130 | 896 | 906 |
| | Log IL12p40 | - | **-.172 (.036) <.001** | -.031 (.04) .438 | .020 (.008) .016 | 892 | 902 |
| | LognIL12p70 | - | **-.085 (.007) <.001** | .003 (.016) .857 | .003 (.003) .343 | 900 | 910 |
| | Log IL13 | - | **-.097 (.01) <.001** | .002 (.015) .908 | .005 (.003) .138 | 898 | 908 |
| | Log TNFα | - | **-.141 (.031) <.001** | -.008 (.045) .864 | .016 (.009) .074 | 894 | 904 |
| | MCP-1 | - | **-.126 (.018) <.001** | -.001 (<.001) .051 | <.001 (<.001) .018 | 912 | 922 |
| **Log QUIN** | Log NFL | **.044 (.011) <.001** | **-.009 (.001) <.001** | .024 (.026) .346 | -.01 (.005) .066 | 534 | 559 |
| | Log GFAP | .027 (.013) .042 | **-.01 (.002) <.001** | .008 (.016) .63 | .002 (.003) .536 | 334 | 356 |
| | Log S100B | **.039 (.011) <.001** | **-.009 (.001) <.001** | -.015 (.026) .575 | -.007 (.005) .163 | 541 | 567 |
| | Log GMCSF | **.042 (.011) <.001** | **-.009 (.001) <.001** | .012 (.007) .067 | <.001 (.001) .961 | 548 | 573 |
| | Log IFNγ | **.037 (.011) .001** | **-.01 (.001) <.001** | .039 (.019) .043 | .002 (.004) .644 | 542 | 568 |
| | Log IL1β | **.04 (.011) <.001** | **-.009 (.001) <.001** | .007 (.018) .709 | -.004 (.004) .318 | 546 | 571 |
| | Log IL1Ra | **.038 (.011) <.001** | **-.009 (.001) <.001** | .026 (.023) .266 | -.006 (.005) .215 | 543 | 568 |
| | Log IL2 | **.039 (.011) <.001** | **-.009 (.001) <.001** | .014 (.012) .26 | -.003 (.003) .283 | 546 | 571 |
| | Log IL4 | **.039 (.011) <.001** | **-.009 (.001) <.001** | -.006 (.012) .631 | -.001 (.002) .514 | 548 | 573 |
| | Log IL5 | **.039 (.011) <.001** | **-.009 (.001) <.001** | .028 (.028) .317 | -.002 (.006) .734 | 544 | 569 |
| | Log IL6 | **.033 (.01) .001** | **-.01 (.001) <.001** | **.079 (.019) <.001** | -.008 (.004) .064 | 525 | 550 |
| | Log IL8 | **.037 (.011) .001** | **-.009 (.001) <.001** | .048 (.031) .127 | <.001 (.006) .98 | 542 | 568 |
| | Log IL10 | **.039 (.011) <.001** | **-.01 (.001) <.001** | .017 (.01) .101 | .002 (.002) .36 | 545 | 571 |
| | Log IL12p40 | **.038 (.011) <.001** | **-.009 (.001) <.001** | .016 (.029) .582 | .008 (.006) .173 | 543 | 568 |
| | Log IL12p70 | **.039 (.011) <.001** | **-.009 (.001) <.001** | -.001 (.012) .918 | <.001 (.002) .952 | 548 | 574 |
| | Log IL13 | **.039 (.011) <.001** | **-.009 (.001) <.001** | .004 (.011) .724 | .002 (.002) .448 | 548 | 573 |
| | Log TNFα | **.038 (.011) <.001** | **-.01 (.001) <.001** | .071 (.035) .043 | .01 (.007) .146 | 539 | 564 |
| | MCP-1 | **.04 (.011) <.001** | **-.009 (.001) <.001** | <.001 (<.001) .284 | <.001 (<.001) .019 | 557 | 582 |

FDR across all fixed effects (time, main cytokine, and interaction): $p<0.001$

**Figure S1: KP metabolites intra-correlations at 2-month post diagnosis**

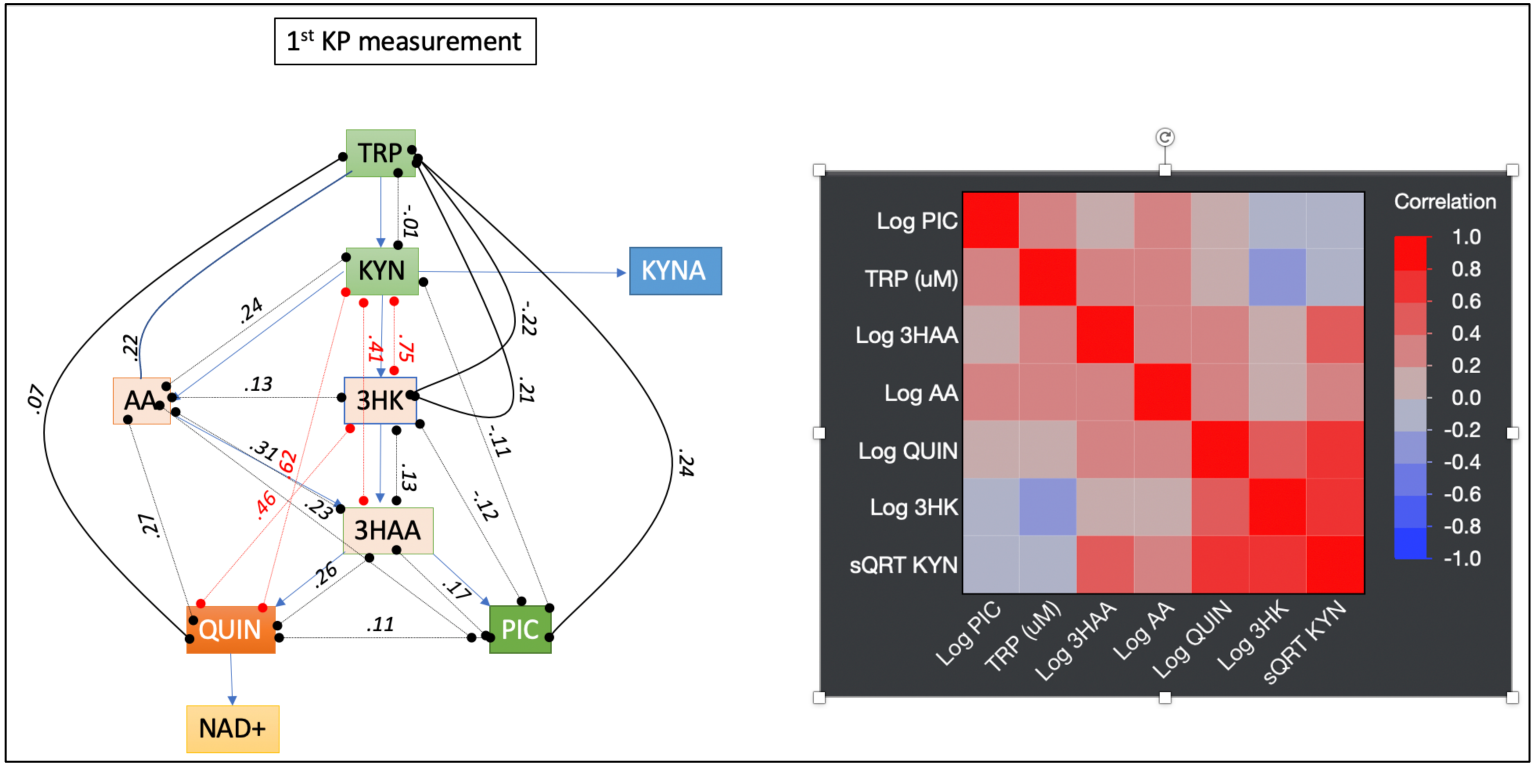

**Figure S2: Extra KP results**

**TRP across the study period**

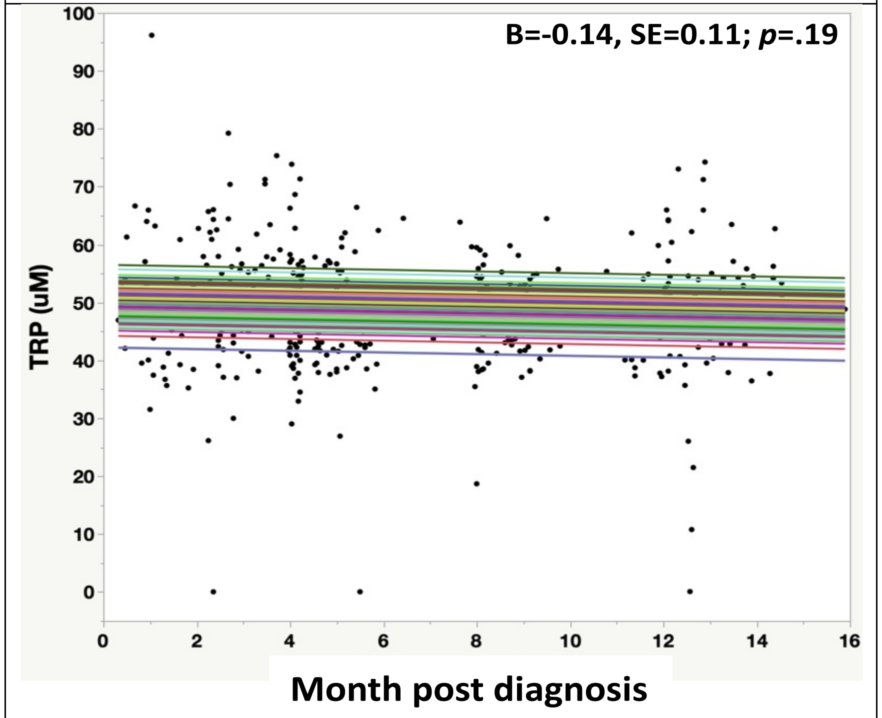


*The KP profile and sex:* We found a main effect of sex where men had higher Log AA compared to women (B=0.18, SE=.087, p=.04) overall, and with greater decrease across time compared to women (B=-0.29, SE=.014, p=.04; Model -2LL=631, AICc=741). Men also had higher Log PIC (B=0.126, SE=.06, p=.04; Model -2LL=310, AICc=320). Finally, men had higher Log QUIN compared to women (B=0.181, SE=.058, p=.002; Model -2LL=369, AICc=379). There was no sex effect for other metabolites.

#### Figure S3: KP and IFN

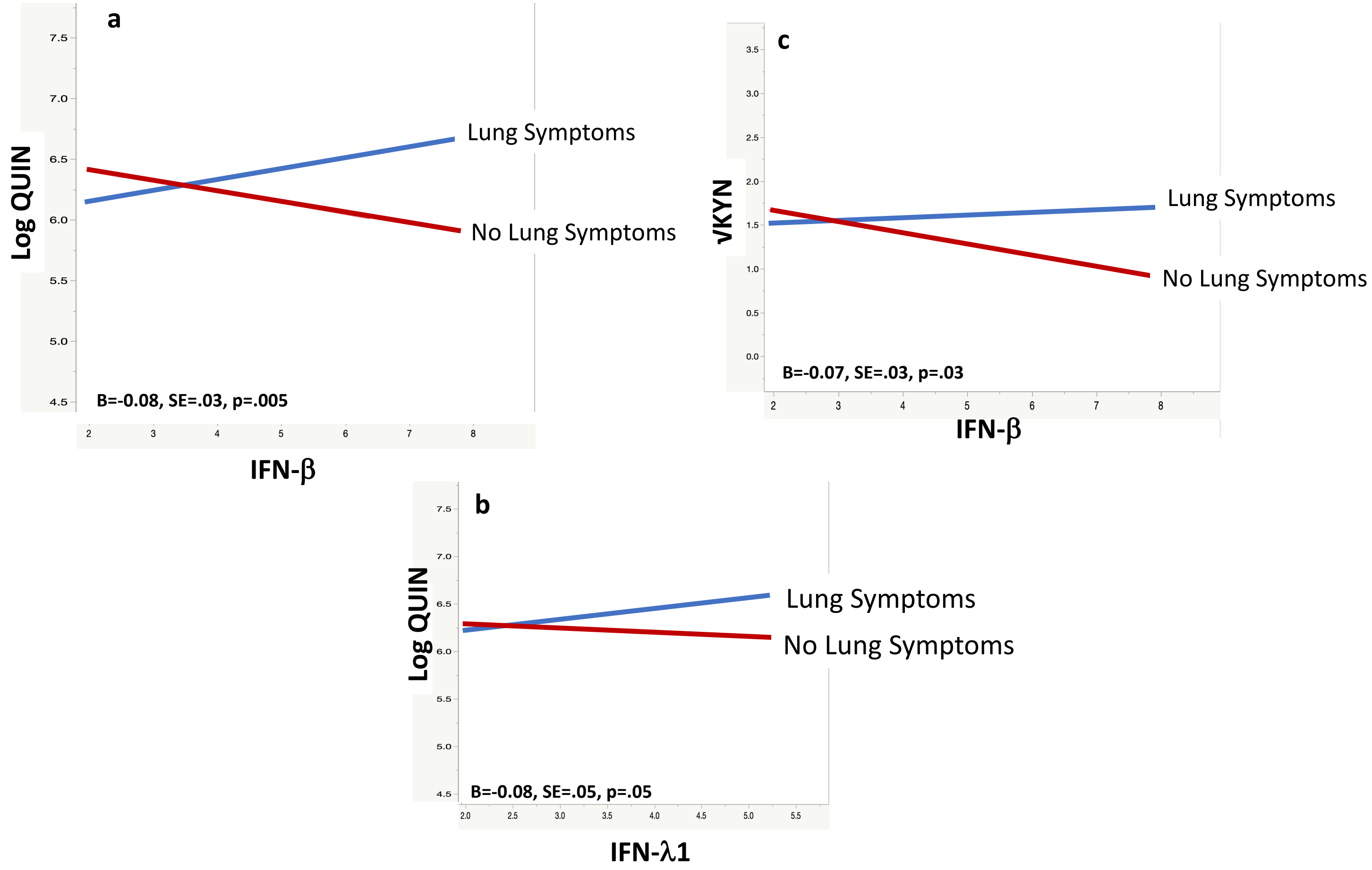


Sub-sample of 62 COVID-19 cases from the same cohort, with half with lung symptoms and half without no lung symptoms. Persistent lung symptoms are defined as persistent chest pain and dyspnoea at 2- and 4- month post diagnosis.

#### Figure S4: Mild to Moderate Anosmia across the study period

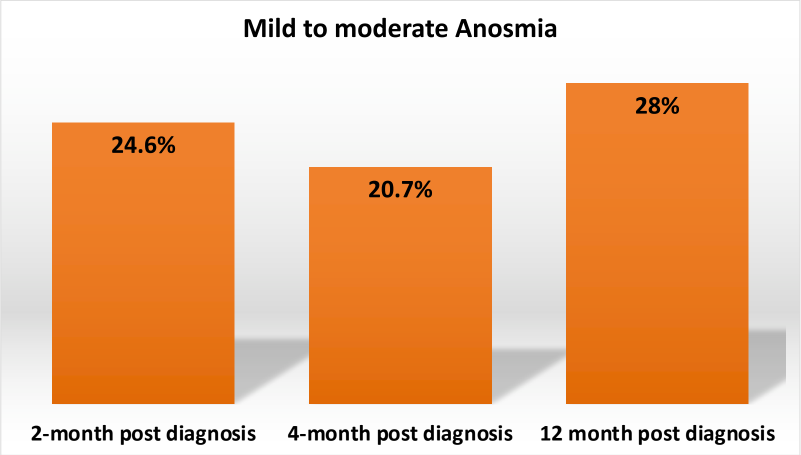


Anosmia was defined as T-score <40 on the NIH Toolbox Odor Identification Test

### Figure S5: Functional status 2- and 4-month post diagnosis

#### 2-month post diagnosis

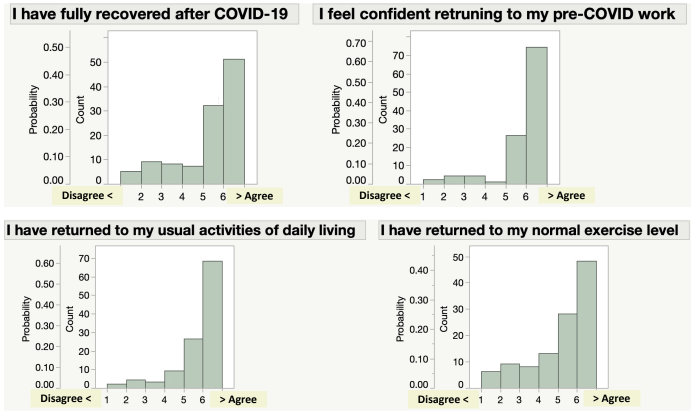


#### 4-month post diagnosis

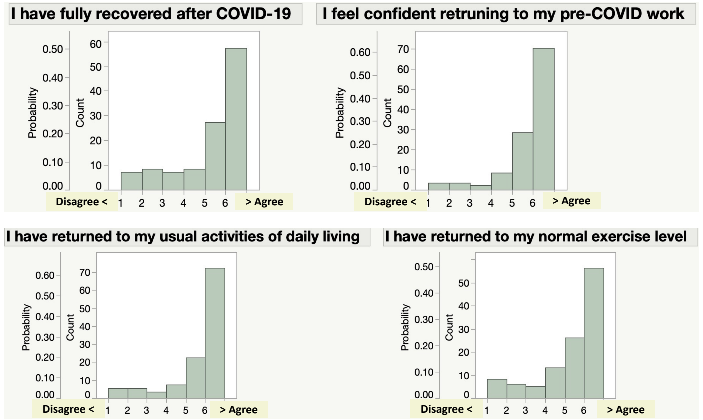
